# Ineffectiveness of international travel restrictions to contain spread of the SARS-CoV-2 Omicron BA.1 variant: a continent-wide laboratory-based observational study from Africa

**DOI:** 10.1101/2024.02.27.24303356

**Authors:** Carlo Fischer, Tongai Gibson Maponga, Anges Yadouleton, Nuro Abílio, Emmanuel Aboce, Praise Adewumi, Pedro Afonso, Jewelna Akorli, Soa Fy Andriamandimby, Latifa Anga, Yvonne Ashong, Mohamed Amine Beloufa, Aicha Bensalem, Richard Birtles, Anicet Luc Magloire Boumba, Freddie Bwanga, Mike Chaponda, Paradzai Chibukira, R Matthew Chico, Justin Chileshe, Wonderful Choga, Gershom Chongwe, Assana Cissé, Fatoumata Cissé, Umberto D’Alessandro, Xavier de Lamballerie, Joana F.M. de Morais, Fawzi Derrar, Ndongo Dia, Youssouf Diarra, Lassina Doumbia, Christian Drosten, Philippe Dussart, Richard Echodu, Tom Luedde, Abdelmajid Eloualid, Ousmane Faye, Torsten Feldt, Anna Frühauf, Simani Gaseitsiwe, Afiwa Halatoko, Pauliana-Vanessa Ilouga, Nalia Ismael, Ronan Jambou, Sheikh Jarju, Antje Kamprad, Ben Katowa, John Kayiwa, Leonard King’wara, Ousmane Koita, Vincent Lacoste, Adamou Lagare, Olfert Landt, Sonia Etenna Lekana-Douki, Jean-Bernard Lekana-Douki, Etuhole Iipumbu, Hugues Loemba, Julius Lutwama, Santou Mamadou, Issaka Maman, Brendon Manyisa, Pedro A. Martinez, Japhet Matoba, Lusia Mhuulu, Andrés Moreira-Soto, Sikhulile Moyo, Judy Mwangi, Nadine Ńdilimabaka, Charity Angella Nassuna, Mamadou Ousmane Ndiath, Emmanuel Nepolo, Richard Njouom, Jalal Nourlil, Steven Ger Nyanjom, Eddy Okoth Odari, Alfred Okeng, Jean Bienvenue Ouoba, Michael Owusu, Irene Owusu Donkor, Karabo Kristen Phadu, Richard Odame Phillips, Wolfgang Preiser, Pierre Roques, Vurayai Ruhanya, Fortune Salah, Sourakatou Salifou, Amadou Alpha Sall, Augustina Angelina Sylverken, Paul Alain Tagnouokam-Ngoupo, Zekiba Tarnagda, Francis Olivier Tchikaya, Noël Tordo, Tafese Beyene Tufa, Jan Felix Drexler

**Author notes:** these authors contributed equally to this work (Carlo Fischer, Tongai Gibson Maponga, Anges Yadouleton).

## Abstract

**Background:** In mid-November 2021, the SARS-CoV-2 Omicron BA.1 variant was detected in Southern Africa, prompting international travel restrictions of unclear effectiveness that exacted a substantial economic toll.

**Methods:** Amidst the BA.1 wave, we tested 13,294 COVID-19 patients in 24 African countries between mid-2021 to early 2022 for BA.1 and Delta variants using real-time reverse transcription-PCR tests. The diagnostic precision of the assays was evaluated by high-throughput sequencing in four countries. The observed BA.1 spread was compared to mobility-based mathematical simulations.

**Findings:** By November-December 2021, BA.1 had replaced the Delta variant in all African sub-regions following a South-North gradient, with a median R_t_ of 2.4 up to 30 days before BA.1 became predominant. PCR-based South-North spread was in agreement with phylogeographic reconstructions relying on 939 SARS-CoV-2 genomes from GISAID. PCR-based reconstructions of country-level BA.1 predominance correlated significantly in time with the emergence of BA.1 genomic sequences on GISAID (p=0.0035, cor=0.70). First BA.1 detections in affluent settings beyond Africa were predicted adequately in time by mobility-based mathematical simulations (p<0.0001). BA.1-infected inbound travelers departing from five continents were identified in five Western countries and one Northern African country by late November/early December 2021, highlighting fast global BA.1 spread aided by international travel.

**Interpretation:** Unilateral travel bans were poorly effective because by the time they came into effect, BA.1 was already widespread in Africa and beyond. PCR-based variant typing combined with mobility-based mathematical modelling can inform rapidly and cost-efficiently on R_t_, spread to inform non-pharmaceutical interventions.

**Funding:** Bill & Melinda Gates Foundation and others.

## Introduction

By December 2023, over 6.9 million people had died from coronavirus disease 2019 (COVID-19) (https://data.who.int/dashboards/covid19/cases). The true number of infections and deaths is probably underreported, particularly in Africa where the diagnostic capacities are low.^1^ In Africa, the World Health Organization (WHO) estimates that only 14% of all Severe acute respiratory syndrome coronavirus 2 (SARS-CoV-2) infections have been detected^2^ and regional post-mortem data suggest the real COVID-19 death toll may be underestimated.^3^

SARS-CoV-2 has evolved rapidly throughout the COVID-19 pandemic. The most pronounced viral change was the emergence of the Omicron variant (BA.1), which was first reported on November 11, 2021, in a patient from South Africa. Within a few weeks, BA.1 was reported in 87 countries,^4^ prevailing over Delta to become the predominant SARS-CoV-2 variant globally by the end of December 2021.^5^ BA.1 had more than 50 non-synonymous mutations compared to ancestral SARS-CoV-2 strains, mostly located in the gene encoding the viral spike protein (https://covariants.org/). These included mutations contributing to effective viral evasion of immune responses elicited by vaccination or prior infection^6^ and mutations favoring BA.1 entry via the receptor-independent endosomal pathway and entailing increased replication of BA.1 in epithelial cells from the upper respiratory tract.^7^ Efficient immune evasion and infection of the upper respiratory tract were likely key to the explosive global spread of BA.1.^8^

In response to the emergence of BA.1, the United States of America, the European Union, the United Kingdom, and several other African and non-African countries restricted international travel by the end of November 2021 for four to six weeks from and to Southern and Eastern African countries, including Botswana, Lesotho, Mozambique, Namibia, South Africa, Swaziland, Zambia, and Zimbabwe.^9,10^ The direct economic loss in South Africa alone caused by these travel restrictions was estimated to be 600 million US dollars.^11^ Despite the unilaterally deployed travel bans, BA.1 spread rapidly to all continents^12^ putting their effectiveness into question. Here, we present the results of an epidemiological molecular study conducted during the BA.1 wave to elucidate the spatiotemporal spread of BA.1 across Africa.

## Methods

### Study participation

We invited 39 laboratories in 34 African countries to join the study. 27 laboratories agreed to participate and were supplied with diagnostic kits (**Fig. 1A**). All laboratories provided results from re-testing specimens from patients or inbound travelers undergoing mandatory PCR testing upon arrival, all of whom were positive for SARS-CoV-2 in routine diagnostic testing.

**Figure 1:**
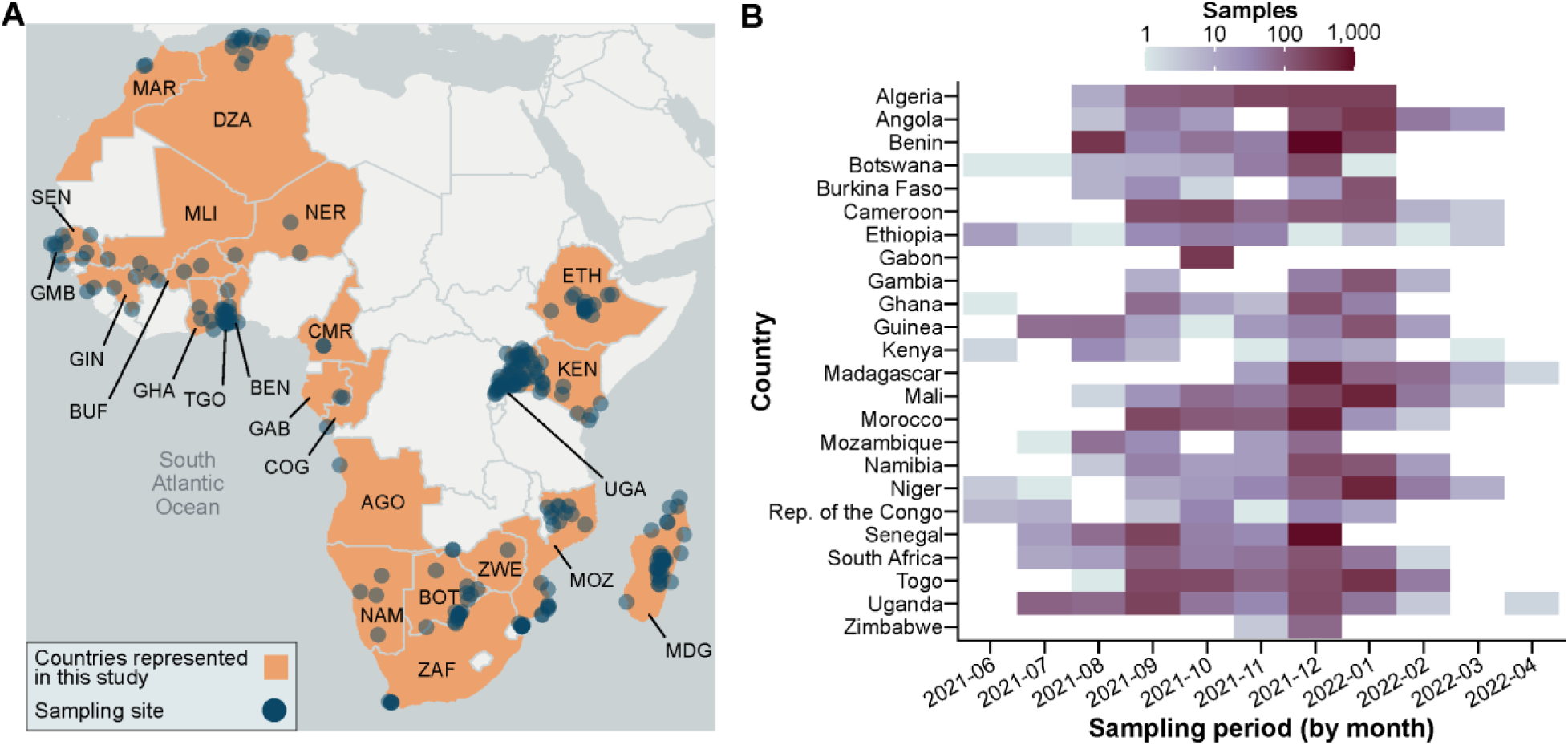
Study setup. (**A**) Geographic distribution of sampling sites and countries (alpha-3 country codes) represented in the study. (**B**) Tested samples among countries by month. The number of tested samples varied between 64 and 1,670 samples per country depending on availability of stored samples and SARS-CoV-2 cases in the participating countries. The geographic presentation of study sites does not imply the expression of any opinion whatsoever concerning the legal status of any country, area, or territory or of its authorities, or concerning the delimitation of its borders. Country abbreviations are given according to alpha-3 ISO codes.

### Molecular testing

A real-time RT-PCR test (**Table S1**) using hydrolysis probes for SARS-CoV-2 detection and genotyping of BA.1 and Delta variants was designed to target a BA.1-specific marker (spike 214 EPE insertion) which is near-absent in other Omicron lineages, and a Delta-specific marker (spike deletion 157/158). The test achieved an in-silico specificity of 98.7% for BA.1 and 99.8% for Delta according to GISAID data available by January 18, 2022 (**Table S2**), when most samples analyzed in this study had been collected (**Fig. 1B**).

### Statistical analyses and data handling

Data cleaning and all analyses were conducted in R version 4.2.1. Patient information included, beyond PCR results, age, sex, location and date of sample collection, laboratory code, and sample ID. Of 14,689 re-tested samples, 13,294 met inclusion criteria (**Table 1**, **S3**).

**Table 1.** Tested samples, typing results and collection period of included samples. Samples with negative or unclear SARS-CoV-2 test, missing collection date or that were sampled before June 2021 or after April 2022 were excluded. Percentage of all reported cases included in this study is shown for the covered collection date range. Samples with missing data on age or sex were included in analyses as both variables were not considered essential. Samples with missing information on collection date, samples that were negative for SARS-CoV-2 on re-testing, samples with unclear results for Delta and BA.1, and samples without unique sample IDs were excluded, as were samples collected before June 2021 as BA.1 was most likely not circulating at that time.

| Country | Total samples | Delta | BA.1 | Other variant | Negative | Unclear | Samples included | Collection date range (dd-mm-yy) | Reported cases | Percentage of reported cases tested |
| --- | --- | --- | --- | --- | --- | --- | --- | --- | --- | --- |
| Algeria | 901 | 733 | 166 | 2 | 0 | 0 | 901 | 18-08-21 – 30-01-22 | 62,111 | 1.5 |
| Angola | 671 | 57 | 471 | 80 | 63 | 0 | 608 | 24-08-21 – 31-03-22 | 53,093 | 1.1 |
| Benin | 1,762 | 269 | 1,234 | 167 | 91 | 1 | 1,670 | 02-08-21 – 27-01-22 | 18,056 | 9.2 |
| Botswana | 231 | 38 | 169 | 24 | 0 | 0 | 231 | 12-06-21 – 16-01-22 | 178,198 | 0.1 |
| Burkina Faso | 197 | 40 | 146 | 11 | 0 | 0 | 197 | 21-08-21 – 28-01-22 | 6,916 | 2.8 |
| Cameroon | 708 | 335 | 220 | 153 | 0 | 0 | 708 | 01-09-21 – 25-03-22 | 36,119 | 2 |
| Ethiopia | 158 | 54 | 12 | 68 | 24 | 0 | 134 | 01-06-21 – 07-03-22 | 197,466 | 0.1 |
| Gabon | 298 | 189 | 0 | 109 | 0 | 0 | 298 | 01-10-21 – 20-10-21 | 5,370 | 5.5 |
| Gambia | 187 | 7 | 174 | 6 | 0 | 0 | 187 | 01-09-21 – 22-02-22 | 2,226 | 8.4 |
| Ghana | 542 | 60 | 145 | 73 | 54 | 210 | 278 | 12-06-21 – 22-01-22 | 60,447 | 0.5 |
| Guinea | 357 | 157 | 188 | 12 | 0 | 0 | 357 | 06-07-21 – 17-02-22 | 12,483 | 2.9 |
| Kenya | 238 | 27 | 15 | 22 | 101 | 73 | 64 | 02-06-21 – 25-03-22 | 152,266 | <0.1 |
| Madagascar | 1,091 | 680 | 130 | 26 | 255 | 0 | 836 | 22-11-21 – 11-04-22 | 20,417 | 4.1 |
| Mali | 1,137 | 187 | 648 | 103 | 198 | 1 | 938 | 02-08-21 – 02-03-22 | 15,804 | 5.9 |
| Morocco | 994 | 551 | 440 | 3 | 0 | 0 | 994 | 01-09-21 – 22-02-22 | 298,636 | 0.3 |
| Mozambique | 210 | 64 | 68 | 45 | 30 | 3 | 177 | 08-07-21 – 11-12-21 | 70,292 | 0.3 |
| Namibia | 486 | 31 | 288 | 75 | 92 | 0 | 394 | 30-08-21 – 23-02-22 | 32,344 | 1.2 |
| Niger | 733 | 46 | 366 | 321 | 0 | 0 | 733 | 27-06-21 – 17-03-22 | 3,310 | 22.1 |
| Republic of the Congo | 95 | 7 | 37 | 51 | 0 | 0 | 95 | 02-06-21 – 12-01-22 | 10,760 | 0.9 |
| Senegal | 1,348 | 410 | 826 | 39 | 72 | 1 | 1,275 | 19-07-21 – 31-12-21 | 22,959 | 5.6 |
| South Africa | 412 | 162 | 239 | 11 | 0 | 0 | 412 | 01-07-21 – 08-02-22 | 1,633,456 | <0.1 |
| Togo | 1,014 | 480 | 533 | 0 | 0 | 1 | 1,013 | 01-08-21 – 22-02-22 | 20,957 | 4.8 |
| Uganda | 823 | 408 | 189 | 115 | 10 | 101 | 712 | 09-07-21 – 14-04-22 | 78,454 | 0.9 |
| Zimbabwe | 96 | 0 | 80 | 2 | 13 | 1 | 82 | 24-11-21 – 13-12-21 | 33,466 | 0.2 |

### Data cleaning for modelling of R_t_ and BA.1 takeover

To reduce the impact of potential false-positive BA.1 PCR test results and of potential BA.1 ancestors harboring the BA.1 marker but not genetically classifiable as BA.1, the dataset was filtered. The dates of first hypothetically expected true BA.1 cases were calculated backwards country-wise considering the date when BA.1 became dominant in our data, the doubling time (set to three days^13^), and population sizes (formula in Supplementary methods). The doubling time was Samples positive for the BA.1 marker that were collected before the estimated first case were removed.

### Estimation of the BA.1 fraction among SARS-CoV-2 infections

First, different generalized linear models (glms) were fitted to the country-stratified PCR data and compared with the moving average of the BA.1 fraction measured by PCR. The moving average was calculated with a window size of 21 days to reduce the impact of sampling gaps. The best model was selected based on the Akaike information criterion (AIC) and optical evaluation of the predicted values. The mean AIC (**Table S4**) and the predicted BA.1 fraction at the beginning of the studied timeframe (June 2021) (**Fig. S1-2**) supported to fit glms using the formula “y ∼ x”. PCR data were grouped by African sub-regions according to the African Union scheme (https://au.int/en/member_states/countryprofiles2). Next, glms were fitted using the grouped and filtered data in R. BA.1 dominance was defined by >50% of tested samples being positive for BA.1.

### Estimation of the time between the first BA.1 case and BA.1 predominance

Countries for which the date of BA.1 becoming the dominant variant was predicted before the first date of BA.1 detection in this study were excluded from calculating the time difference between the first BA.1 case and BA.1 predominance.

### R_t_ estimation

To estimate R_t_ for BA.1, a glm was employed to estimate the relative fraction of BA.1 among all SARS-CoV-2 cases for each country represented in this study based on PCR results. Daily estimated BA.1 cases were calculated by multiplying the modelled BA.1 fraction with reported SARS-CoV-2 cases. R_t_ was calculated using the EpiEstim package in R. The serial interval was set to 3.3 days (standard deviation 2.4 days) based on epidemiological analyses of BA.1.^8,13^

### Estimation of daily BA.1 and Delta variant cases

To estimate daily infections with the BA.1 and Delta variants, the estimated fraction of a specific variant among all SARS-CoV-2 cases in one country was multiplied with the country-specific smoothed new cases per million people as reported by WHO (accessed via Our World in Data by May 20, 2022).

### Simulation of the BA.1 spread

The global spread of BA.1 was simulated using a Global Epidemic and Mobility Model (GLEAM) in the GLEAMviz Simulator version 7.2^14^. The simulator combines a SEIR (“Susceptible”, “Exposed”, “Infected” and “Recovered”) model (details in **Fig. S3**) with a metapopulation network approach assigning cells to closest airports. Roughly 3,300 subpopulations are interconnected by flight connections (retrieved from Official Airline Guide and International Air Transport Association, https://www.iata.org). Commuting data is included to simulate short distance spread. Multi-run simulations (ten simulations) were conducted for each model. Simulations were started on November 11, 2021, corresponding to the collection date of the first identified BA.1 cases in Botswana and simulated for 109 days until end of February, 2022, when BA.1 cases declined.^15^ Active BA.1 cases at simulation start were defined for Cape Town, Durban, Johannesburg (South Africa), and Gaborone (Botswana) where first BA.1 cases were identified.^16^ The number of BA.1 cases for the simulation were calculated by multiplying the PCR-based predicted BA.1 fraction for South Africa and Botswana with reported SARS-CoV-2 cases. As SARS-CoV-2 cases are likely underreported,^2^ 11 additional simulations were calculated with 1.5-fold increasing BA.1 starting cases (**Table S5**). Each simulation was calculated with 13, 31, and 49% pre-existing immunity based on recent observations,^17^ resulting in 36 simulations.

The GLEAMviz simulation of daily BA.1 cases fitting best the estimated daily BA.1 cases was identified by analyzing least sum of squares of data grouped by African regions. Correlation was analyzed by Pearson correlation coefficient.

### Phylogeographic analyses

For phylogeographic analyses, all African GISAID entries classified as BA.1 available on February 26, 2023, with a collection date before December 2021 were retrieved and filtered for at least 95% of genome coverage. A discrete phylogeographic diffusion model was applied in BEAST using a tip-dated dataset and country-level midpoints to identify signs of geographical migration as previously described.^18^ SpreaD3 was applied to visualize the geographical association of phylogenetic lineages and geographical diffusion. In addition to posterior probabilities of geographic transitions, Bayes factors were calculated by dividing the likelihood of the alternative hypothesis (origin in South/Southern Africa) by the likelihood of the null hypothesis (origin not in South/Southern Africa).

### High-throughput sequencing

In Benin, SARS-CoV-2 samples were prepared for HTS-based sequencing using the EasySeq™ SARS-CoV-2 WGS Library Prep Kit (NimaGen). Sequencing was conducted using the MiSeq Reagent Kit v3 (Illumina). Up to 80 samples prepared using different adapters, were pooled in one sequencing run. HTS reads were analyzed using the CoVpipe pipeline (https://gitlab.com/RKIBioinformaticsPipelines/ncov_minipipe/-/tree/master/covpipe). In South Africa and Botswana, samples were sequenced using an Oxford Nanopore Technologies-based workflow as described previously.^19^ In Guinea, samples were sequenced using both Illumina NextSeq500 system and Oxford Nanopore Technologies.

## Results

The clinical sensitivity and specificity of the provided genotyping PCR kits among 811 samples from this study were 92.4% and 100% for the Delta test and 99.6% and 97.6% for the BA.1 test in reference to HTS-based classification of SARS-CoV-2 sequences (**Table S6**). Eight samples wrongly reported as BA.1 by laboratories with available HTS data all lacked the spike 214 EPE insertion targeted by the test, suggesting false-positive results during reporting or BA.1 contamination of samples which is supported by high Ct values (>35) in the BA.1 test for four samples where Ct values were available. The predicted high specificity of BA.1 detection according to genomic database entries was confirmed *in vitro* by the absence of the BA.1 marker in 545 SARS-CoV-2-positive respiratory samples from Benin, Western Africa, collected between January and April 2021.^20^ In total, 13,294 samples from laboratory-confirmed COVID-19 patients from 24 African countries and 225 municipalities (**Fig. 1A**) sampled during mid-2021 to early 2022 were included in this study (**Fig. 1B**).

Across African countries, BA.1 replaced Delta as the predominant SARS-CoV-2 variant by December 2021 (**Fig. 2A**, Delta fraction and **2B**, BA.1 fraction; **Fig. S4**). 33 samples collected before November 2021 in Benin, Ghana, Mali, Niger, and Uganda were tested positive for BA.1 but could not be validated by full genome sequencing due to exhaustion of clinical materials and potential contamination during sample or library preparation prior to HTS.^21^ Although near-parallel detection of BA.1 marker-positive samples in five African countries may hypothetically represent emergence of ancestral SARS-CoV-2 strains prior to BA.1 emergence, those samples were excluded from downstream analyses to minimize the impact of potentially false-positive samples. Moreover, data were analyzed based on time points when BA.1 became dominant and modelled early cases backwards to further increase the robustness of downstream epidemiological analyses. Comparing continent-wide PCR data, BA.1 became the dominant variant (>50% detection) on November 8 (95% CI, −3/+3 days) in Southern Africa, December 1 (95% CI, −1/+1 days) in Western Africa, December 10 (95% CI, −3/+3 days) in Central Africa, December 13 (95% CI, −3/+3 days) in continental Eastern Africa, and December 25 (95% CI, −1/+1 days) in Northern Africa (**Fig. 2C**; **Fig. S4**). These data suggest that BA.1 was widely spread in and beyond Southern Africa when travel restrictions were put into place within and beyond Africa (**Fig. 2C**). Not removing potentially false-positive BA.1 samples had a minimal effect on the predicted BA.1 spread, changing the dates of BA.1 dominance for Central, Eastern, and Northern Africa by one day each, and for Western Africa by two days, highlighting the robustness of our approach (**Table S7**). The South-North gradient suggested by those data is consistent with the emergence of BA.1 in Southern Africa and phylogeographic reconstructions based on available GISAID sequences collected between June 2021 and March 2022 (**Fig. 3**). Those phylogeographic reconstructions suggested transitions from South Africa to all African regions, while only one backwards transition was reconstructed at high posterior probability from Nigeria to South Africa in late December 2021. A South African or Southern African origin of BA.1 was reconstructed at very strong (Bayes factor 44.5) and moderate (Bayes factor 4.4) support, respectively. Delayed BA.1 introduction into Northern Africa may be associated with reduced land connectivity imposed by the Sahara Desert^22^ or by the lack of regional BA.1 cases when travel bans were implemented.^10^ Similarly, border closure in Madagascar until late 2021 delayed BA.1 introduction until January 2022 (**Table S8**). Across African countries, the median time between the first BA.1 detection and BA.1 predominance was 19 days (95% CI, 10-99.0) (**Fig. 4A**; **Fig. S4**), which was comparable to BA.1 spread in high-income countries.^5,23^ Combining all country-level PCR data from Africa, the BA.1 effective reproduction number R_t_ was 2.4 during the 30 days until BA.1 became the dominant variant (**Fig. 4B**; **Fig. S4**; **Table S9**), which was lower than an overall R_0_ of 3.7 (95% CI, 3.3–4.1) reconstructed for SARS-CoV-2 variants in general in Africa^24^ and an average R_t_ of 3.4 for BA.1 among countries in Africa, the Americas, Asia and Europe.^25^ The relatively low R_t_ may be a consequence of underreporting particularly in times of high case numbers which would also explain the observed decrease of R_t_ when cases increased, or by different serial interval estimations which vary between 3.0 and 5.5 in the literature.^8,24^ In the combined country-level PCR data, R_t_ dropped below one within 40 days after BA.1 became dominant, likely due to widespread immunity following explosive BA.1 spread.^26^ This interpretation is in line with the steep increase of reported cases likely corresponding to short duration of the BA.1 wave in Africa (**Fig. S5**).^12,26^ Retrospective analyses of >67,800 SARS-CoV-2 genomes available in GISAID by mid-2023 revealed that within one week after the new BA.1 variant was first reported to the WHO on November 24, 2021, BA.1 had spread to all African regions except Northern Africa (**Fig. 5A-E**; blue color, BA.1 submission date; pink color, sample collection date; **Fig. S6**). Genomic sequence and PCR-based data from this study were thus consistent with the widespread early occurrence of BA.1 across African regions.

**Figure 2:**
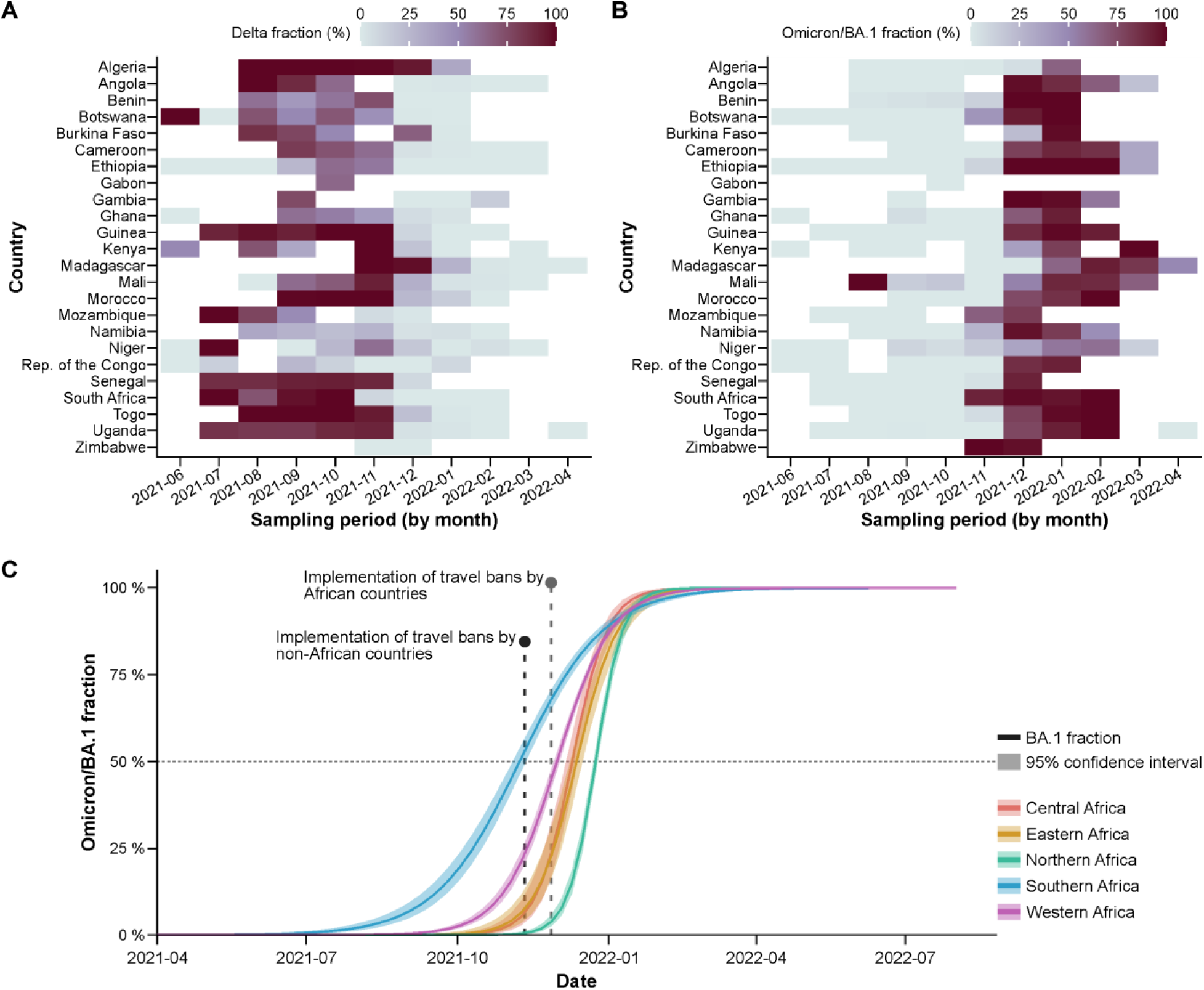
Epidemiology of Omicron/BA.1 in Africa. (**A**) Fraction of samples positive for the Delta marker. (**B**) Fraction positive for the BA.1 marker. (**C**) Modelled increase in BA.1 fraction of all SARS-CoV-2 infections per African region based on PCR testing. All analyses were repeated excluding travelers to confirm robustness (**Fig. S2**). Central and Eastern Africa curves for BA.1 fraction increase overlap. Not-removing early BA.1 cases had minimal effects changing the date of BA.1 dominance for Central, Eastern, and Northern Africa by one day each, and for Western Africa by two days. The date of BA.1 dominance for Southern Africa was not affected.

**Figure 3:**
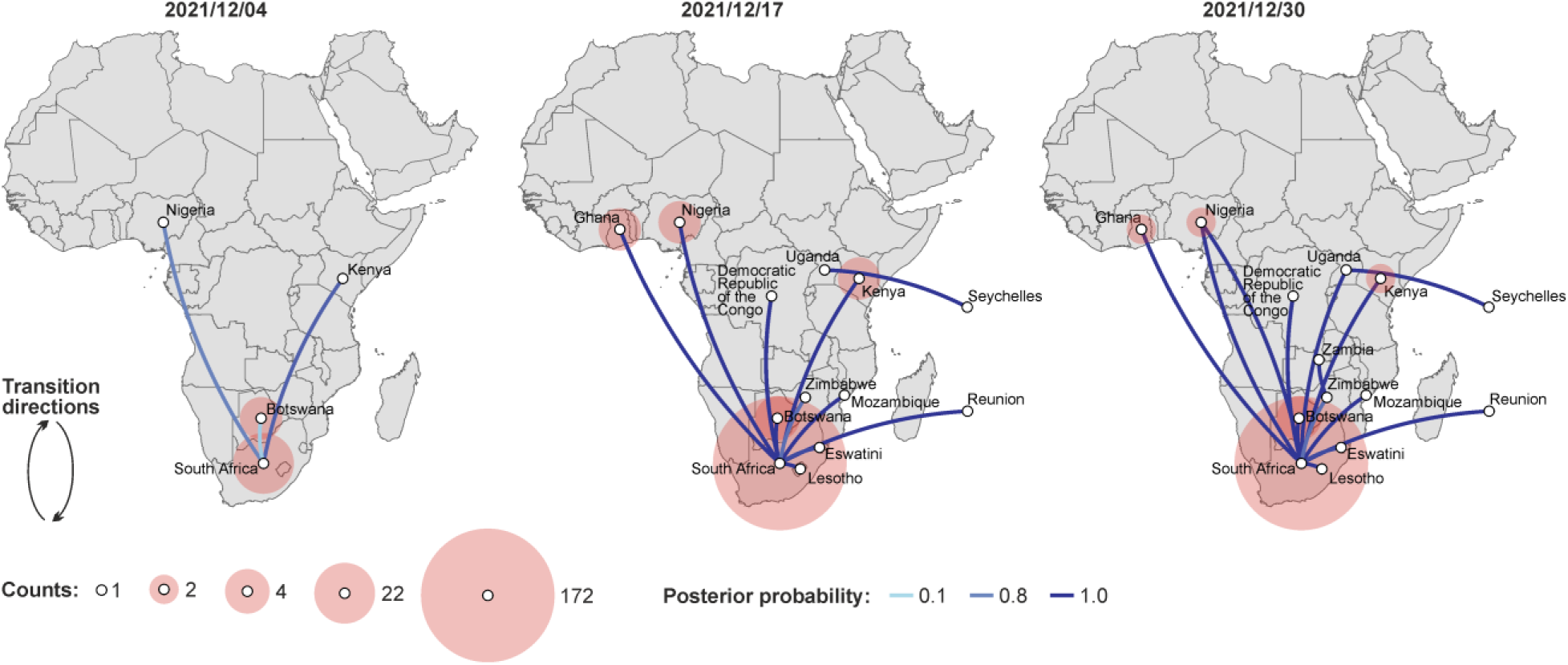
Phylogeographic analyses SARS-CoV-2 Omicron/BA.1 spread in Africa. African BA.1 sequences available on GISAID were filtered by at least 95% genome coverage and sequences with potentially false collection date (to early) were removed resulting in 939 analyzed genomes. Transitions and reward counts are shown for three timepoints. Only completed transitions are shown. Bayes factors above 3 supported transitions between South Africa and Botswana, Ghana, Kenya, Mozambique, Nigeria (**Fig. S6**).

**Figure 4:**
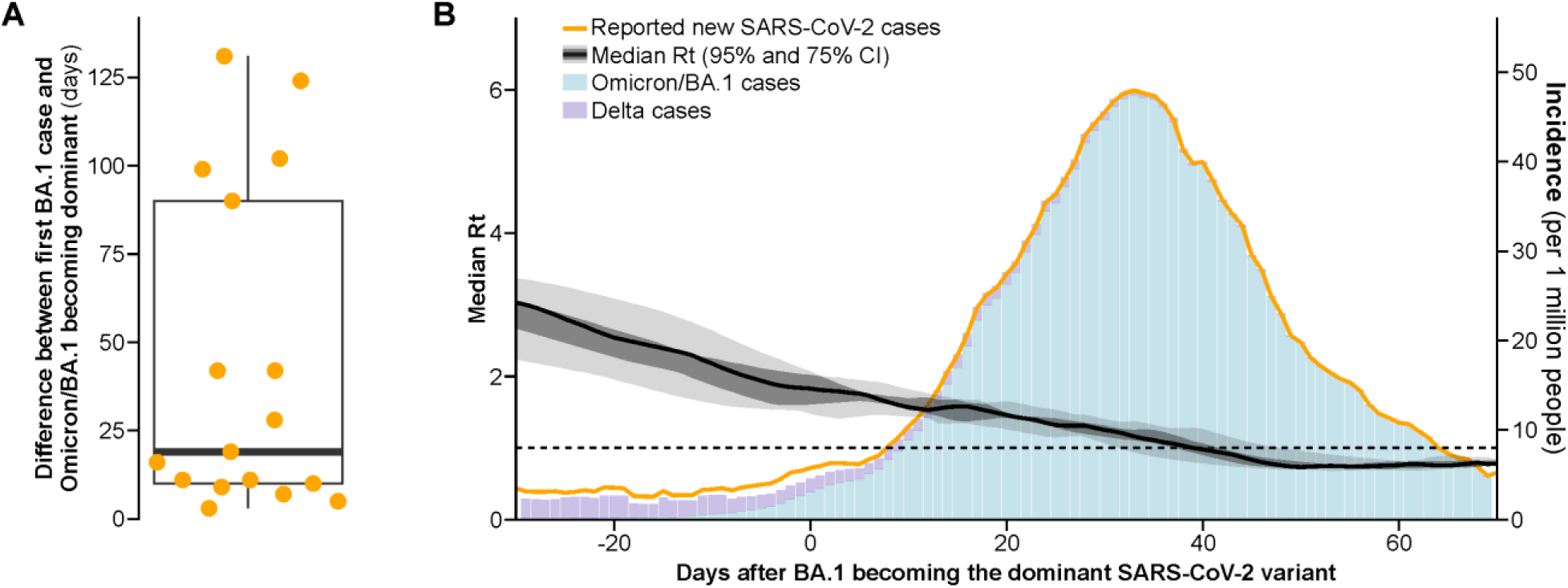
Speed of Omicron/BA.1 spread. (**A**) Days until BA.1 became the dominant SARS-CoV-2 variant after its first detection by PCR. (**B**) Smoothed R_t_ and the incidence among countries represented in this study. All analyses were repeated excluding travelers to confirm robustness (**Fig. S4**).

**Figure 5:**
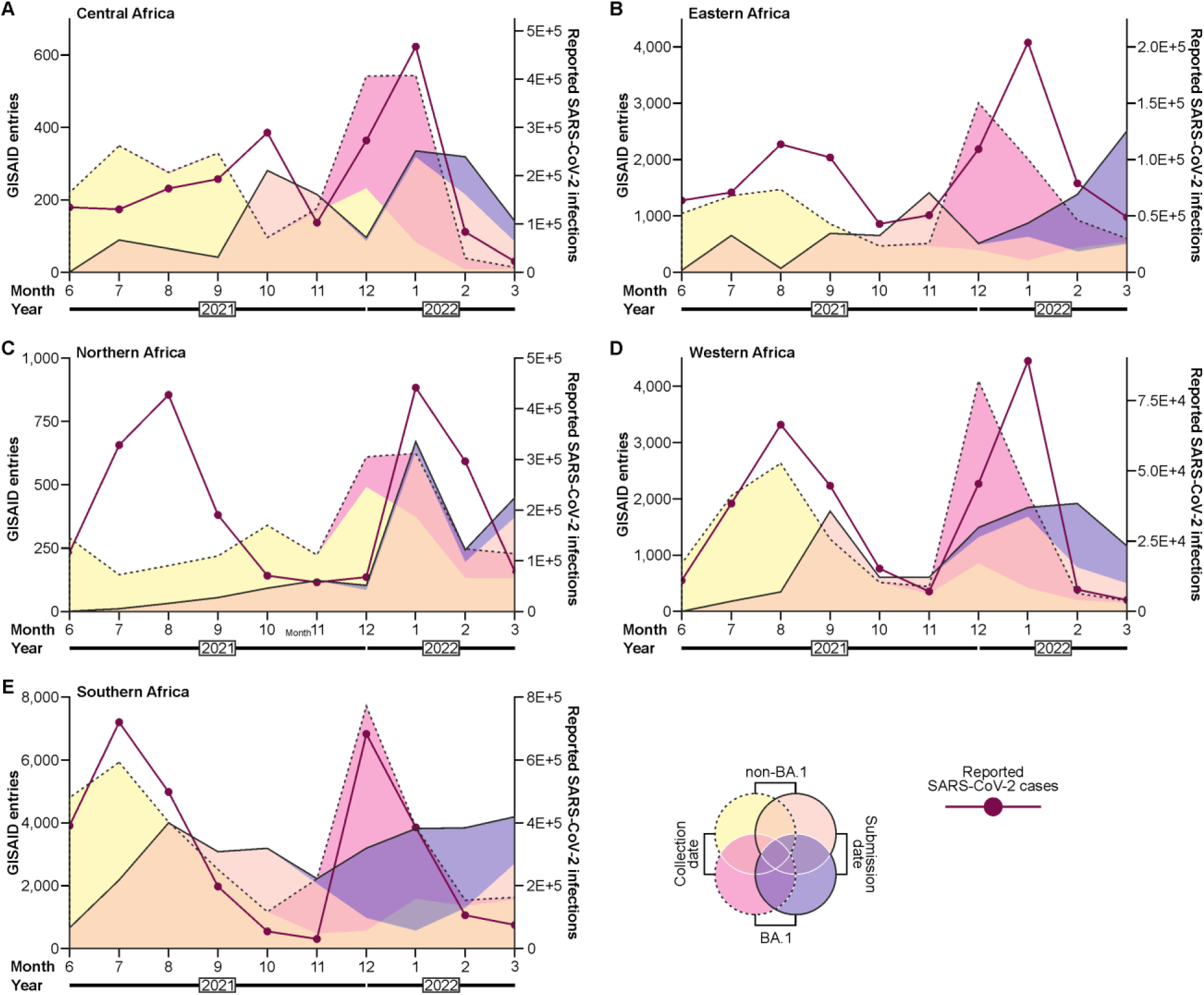
SARS-CoV-2 sequencing and genome submissions. BA.1 and non-BA.1 sequences submitted to GISAID by collection and submission date from (**A**) Central Africa, (**B**) Eastern Africa, (**C**) Northern Africa, (**D**) Western Africa, (**E**) Southern Africa.

The fast spread of SARS-CoV-2 is known to be facilitated by human mobility, including both short- and long-distance travel.^27^ The importance of long-distance travel for the spread of BA.1 was consistent with the detection of BA.1 among inbound travelers in our study. Considering ten countries for which information on the testing of travelers was available, inbound travelers departing from Burkina Faso, Nigeria, and Mauritania were tested positive for BA.1 in Senegal and Togo by November 24, 2021, when the new variant first was reported to WHO. Two weeks later, inbound travelers were tested positive for BA.1 in another four countries (Algeria, Niger, Senegal, Togo) (**Table S10**). Travelers tested BA.1 positive before December 8, 2021, departed from diverse locations in five continents (**Table S10**), highlighting fast global spread of BA.1 and suggesting a BA.1 emergence several weeks before its first detection. This interpretation was in line with GISAID data, retrospective analyses of BA.1 genomes from England^15^, and early phylogenetic analyses reconstructing the most recent common ancestor of BA.1 to early October 2021.^4^

To evaluate the predictability of the BA.1 spread using a ready-to-use user interface based simulation tool, a SEIR global epidemic and mobility model was simulated using the GLEAMviz online tool, which has been used to study the global dispersion of SARS-CoV-2.^27^ The simulation was calculated with PCR-informed starting cases and 1.5-fold increasing starting cases to consider potential underreporting.^2^ Pre-existing immunity was set in a range of 13-49% according to the observed cross-protection from BA.1 infection by a previous SARS-CoV-2 infection (**Table S5**).^17^ The overall best accordance between simulated BA.1 infections and PCR-based estimated BA.1 cases was observed in the simulation assuming 86.5-fold increased starting cases and pre-existing immunity of 13% compared to the original PCR-informed model (**Fig. 6**). In this SEIR simulation, one BA.1 case in 100,000 inhabitants was first simulated in Southern Africa (by November 29, 2021), 17 days later in Eastern Africa, 23 days later in Central Africa, and roughly one month later in Western and Northern Africa (**Table S11**). The modeled South-North gradient concurred with our continent-wide PCR data (**Fig. 2C**). Conversely, the simulation deviated from PCR-based data and GISAID data regarding the emergence of BA.1 in Eastern Africa (later in PCR and GISAID than in model data) and Western Africa (earlier in PCR and GISAID than in model data) (**Fig. 7A-B; Table S8**). These discrepancies may be a consequence of mobility restrictions affecting more Eastern than Western Africa and early introduction of BA.1 into West Africa by long-distance travel as suggested by our PCR and phylogeographic data (**Fig. 3; Fig. S7**). Overall, the simulated BA.1 spread was clearly slower than the spread according to case data. Relatively slower simulated spread may be due to an underestimation of the BA.1 cases at the simulation start, which is in line with early BA.1 detection outside of Southern Africa in both our molecular data and GISAID entries. This simulation was significantly correlated with both PCR-based estimated cases (p = 0.0414, cor = 0.53) and with GISAID entries (p < 0.0001, cor = 0.58) (**Fig. 7C-D**). Moreover, the simulated BA.1 introduction date in non-African countries was significantly earlier in countries which reported BA.1 cases by mid-December 2021 when first estimates of global BA.1 spread had become available (https://www.ecdc.europa.eu/en/news-events/epidemiological-update-omicron-data-29-november-2021) than in countries that did not report early BA.1 cases (Kruskal-Wallis test, p < 0.0001) (**Figure S8**). The correlation of simulated and observed BA.1 introductions thus supported the overall usefulness of mobility-based simulations to predict global spread of an emerging SARS-CoV-2 variant, particularly to identify countries of highest importation risk.

**Figure 6:**
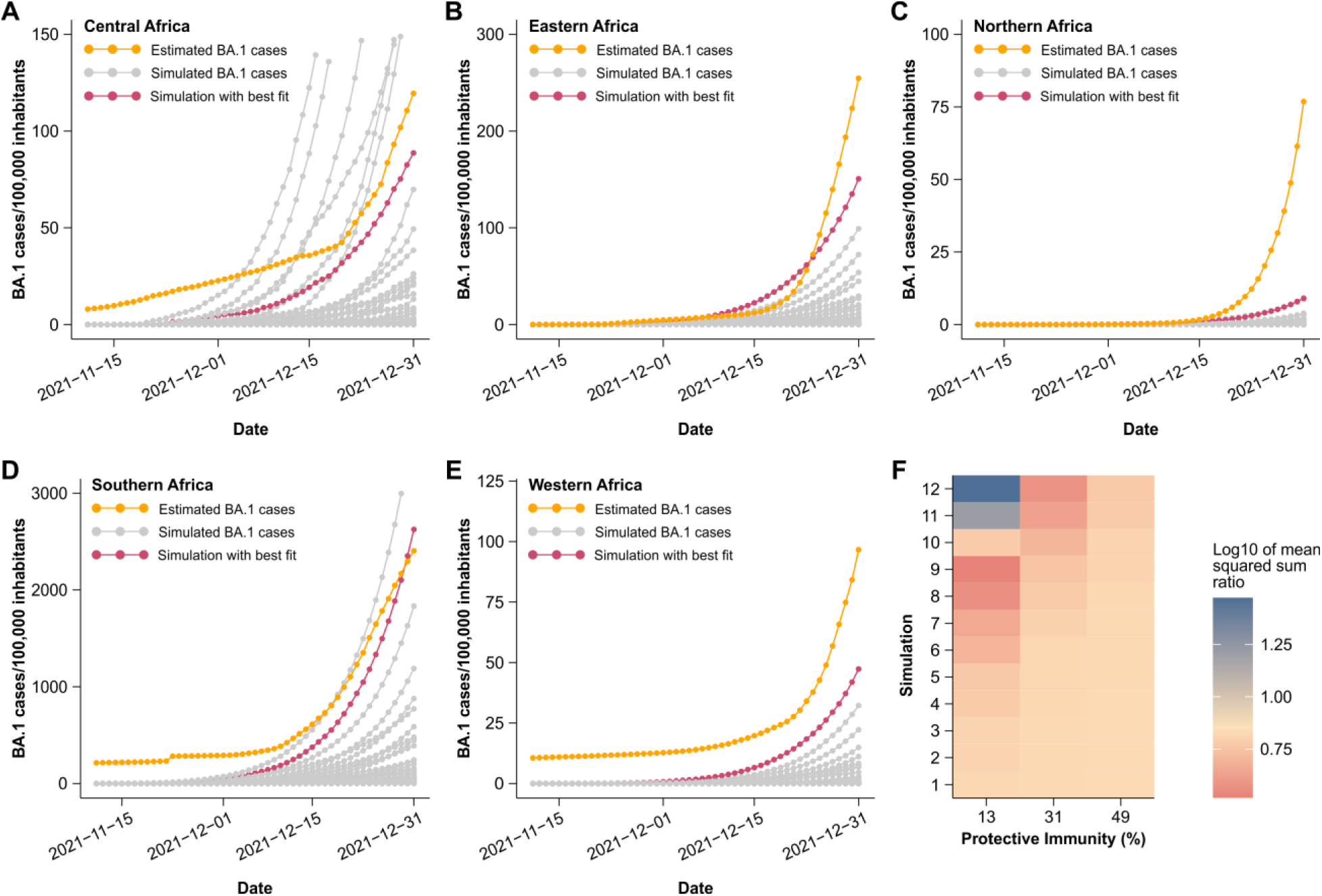
Simulation of Omicron/BA.1 cases. Comparison of estimated and simulated BA.1 cases for Central Africa (**A**), Eastern Africa (**B**), Northern Africa (**C**), Southern Africa (**D**), and Western Africa (**E**). (**F**) Average fit of simulated BA.1 cases with BA.1 cases estimated from PCR results and reported SARS-CoV-2 cases. Concordance of estimated BA.1 cases and simulated cases was determined by least sum of squares over all African regions.

**Figure 7:**
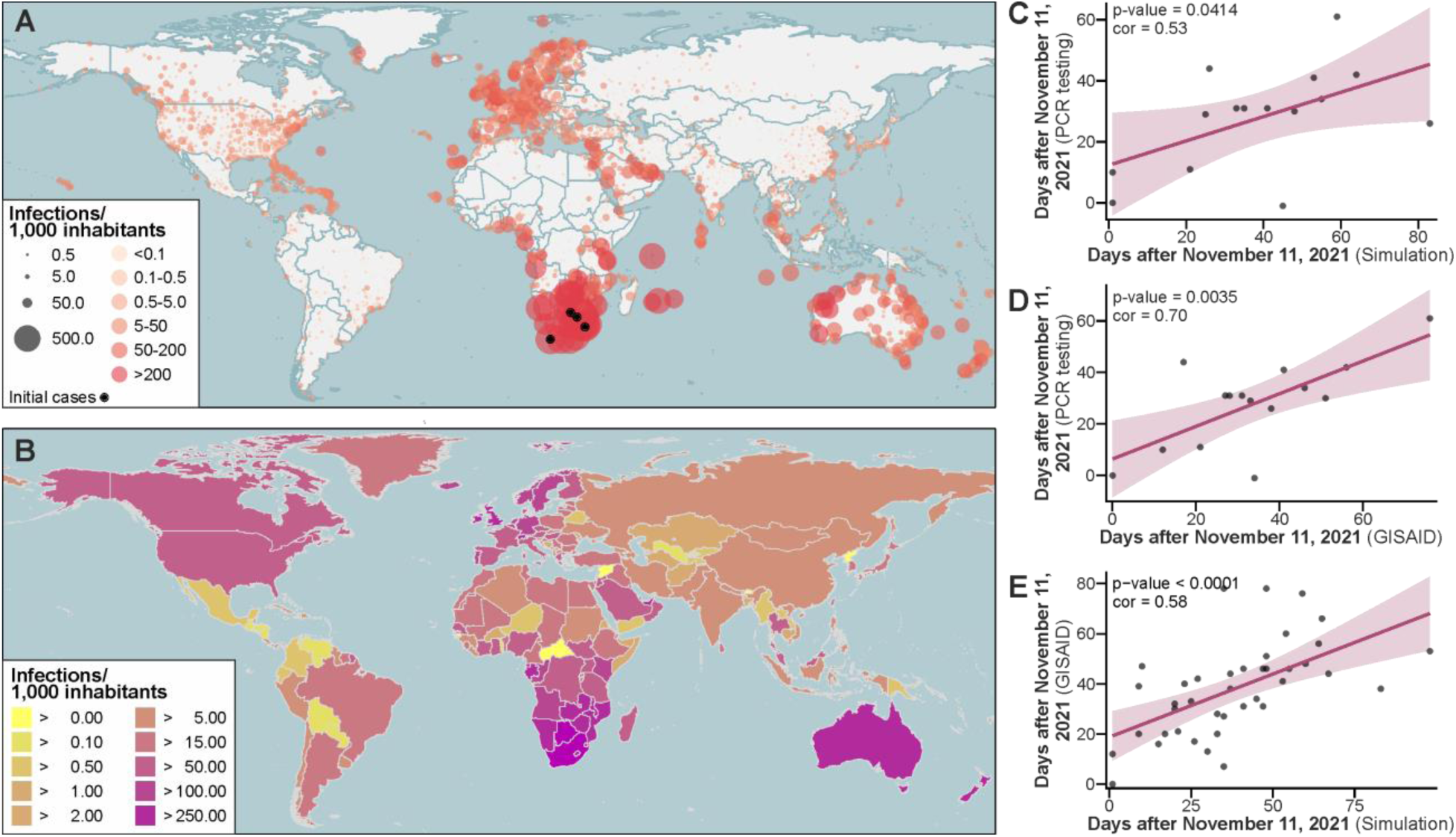
Simulated global spread of BA.1 and correlation with molecular testing. Simulated BA.1 infections by the end of February 2022 on municipality (**B**) and country level (**B**). Defined initial cases are shown by black dots. See **Fig. S1** for simulation setups. (**C**) Correlation of days following November 11, 2021, when BA.1 became the dominant SARS-CoV-2 variant in models based on our PCR testing and when one BA.1 infection was simulated in 100,000 inhabitants using GLEAMviz. Only countries in which the increase of the predicted BA.1 fraction from 2 to 64% (5 doubling intervals) took between 7.5 (half of expected time considering 3 days doubling time) and 90 days (6-times expected days) were considered in C and D to reduce the effect of non-representative data. Madagascar was excluded from the analyses due to strict border closures delaying BA.1 introduction. All values in C-E are on country level. All correlations in C-E were calculated by Pearson correlation. (**D**) Correlation of days following November 11, 2021, when BA.1 became the dominant SARS-CoV-2 variant in models based on our PCR testing and when at least 10 BA.1 sequences were deposited in GISAID (collection date). (**E**) Correlation of days following November 11, 2021, when 1 BA.1 infection was simulated in 1,000 inhabitants using GLEAMviz and when at least 10 BA.1 sequences were deposited in GISAID (collection date).

Surveillance of SARS-CoV-2 variants is commonly done by HTS-based full genome generation of selected samples. However, this approach is relatively costly and time-consuming. Considering standard procedures and products, typing a single sample was roughly six times more expensive and three times more time-consuming by HTS which also requires bioinformatics (USD 49.86, 5.6 minutes) compared to PCR-based typing which only requires laboratory personnel (USD 7.82, 1.9 minutes) (**Table S12**). Moreover, HTS-based sequencing requires access to technical infrastructure which is often limited, resulting in delays between sample collection and availability of sequencing results.^28^ A time-based analysis of 67,821 GISAID entries showed that the median time between sample collection and sequence submission to GISAID ranged between 38 and 132 days for the African regions, irrespective of whether BA.1 or non-BA.1 entries were deposited (**Fig. 8**). Additional to technical reasons, the comparative time-stamped analysis highlighted that neither African countries, nor supranational organizations reported BA.1 sequences with delay, which may hypothetically have been observed in consequence of fear of unilateral travel restrictions.

**Figure 8:**
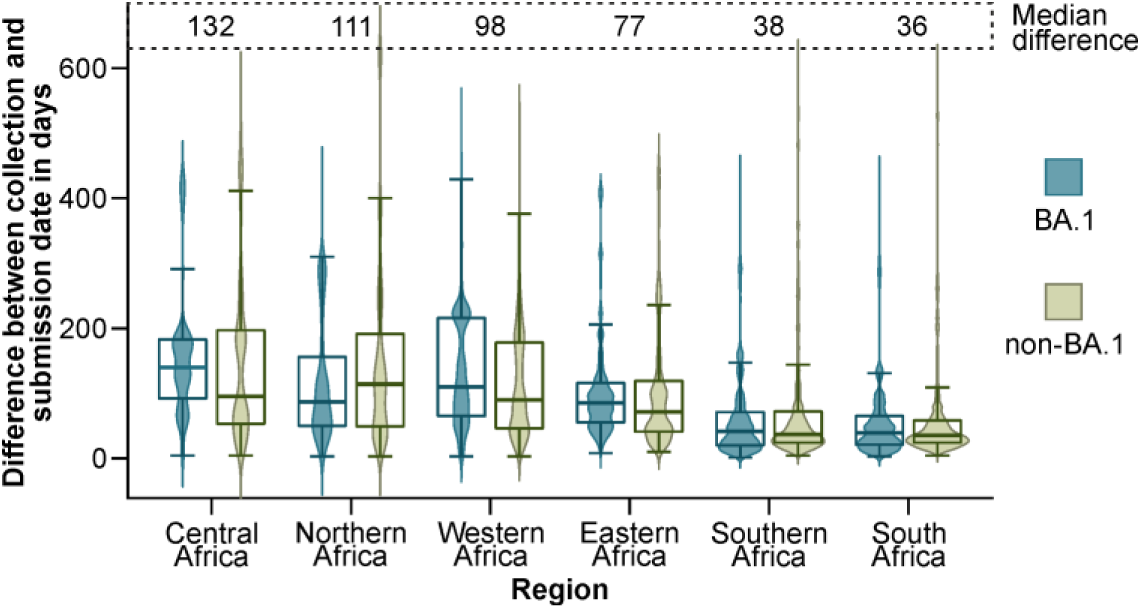
Difference between collection and submission date for SARS-CoV-2 entries from African countries in GISAID. In total, 67,821 GISAID entries were analyzed including 23,076 BA.1 entries.

## Discussion

Our study provides strong evidence that BA.1 spread rapidly across Africa and beyond before travel restrictions were implemented for South and East African countries. Our continent-wide data thus objectively demonstrate the ineffectiveness of travel bans in the context of insufficient surveillance. Upon the emergence of any new SARS-CoV-2 variant, understanding its pathogenicity, pace of transmission, its ability to evade pre-existing immunity, and its spatial distribution are crucial elements. For all four factors the ability to identify a new variant is pivotal. Sequencing 0.5% of all cases within 21 days after sample collection provides a good chance to detect new SARS-CoV-2 variants efficiently and early.^28^ However, establishing efficient genomic surveillance infrastructures is a major economical and organizational challenge. Despite globally increasing sequencing capacities, only 5% of low- and middle-income countries have reached this benchmark and some African countries still rely on external capacities for genome sequencing and submission.^28^ Moreover, SARS-CoV-2 infections are probably widely underreported in Africa,^2^ causing sampling biases which may also affect genomic surveillance. To allow for efficient interventions^29^ strengthening and harmonizing surveillance systems on a supranational level, establishing strategic sampling frameworks, and supporting the sharing of surveillance data is hence critial.^30^ We demonstrate that specifically designed real-time RT-PCR tests have the potential to strengthen the surveillance of newly identified SARS-CoV-2 variants as a complement to sequencing before sequencing can be scaled up on a continent-wide scale. Importantly, molecular typing assays such as the one we used may be most valuable immediately after variant emergence because tested markers may emerge convergently in other than the target lineage.^20^

Our findings make future preparedness in resource-limited settings conceivable at four levels. First, sequencing capacities including HTS- and PCR-based protocols should be upscaled to allow early detection of new variants. Second, upon the emergence of a new variant, real-time RT-PCR assays for variant typing need to be designed, validated, produced, and provided rapidly across the entire region. Third, based on the typing results, mobility-based simulations can be applied to predict countries of highest importation risk by land or air travel. The identification of these countries is essential to reduce the risk of dispersion from secondary hubs. Global spread of BA.1 occurred predominantly from affluent settings outside of Africa with intense flight connectivity, such as the United States of America.^31^ Importantly, such mobility-based simulations are dependent on regularly updated mobility information and may require continuous funding by supranational actors such as WHO. Finally, the typing and simulation results can serve as basis to implement non-pharmaceutical interventions including travel restrictions or border closures based on evidence.^27,32^ Our study is limited by heterogeneous sampling in time and space and by lack of SARS-CoV-2 genomic data from all BA.1 PCR-positive patients. However, the PCR test was exhaustively validated in four countries and geographically widespread testing substantiates robustness of our findings.

In conclusion, our results highlight that unilaterally implemented travel restrictions failed to contain BA.1. We demonstrate how PCR-based variant typing allowed us to assess the spatiotemporal spread of a new SARS-CoV-2 variant rapidly and economically on a continent-wide scale, contributing to the design of containment strategies for future SARS-CoV-2 variants and other emerging pathogens.

## Data Availability

Respiratory samples sent to laboratories for COVID-19 testing by attending physicians were re-tested in an anonymized fashion using the COVID-19 test developed in this study. Ethical approval for re-testing and scientific usage was provided by the institutional research ethics board (IRB) of Charite-Universitaetsmedizin Berlin (EA2/028/22) and by IRBs from Burkina Faso, Laboratoire National de Reference-Grippes (2020-7-126), Cameroon, Centre Pasteur du Cameroun (2020/05/1224/CE/CNERSH/SP), Ghana, Kumasi Centre for Collaborative Research in Tropical Medicine (KCCR), KNUST (CHRPE/AP/566/21), Kenya, Jomo Kenyatta University of Agriculture and Technology, Department of Biochemistry (JKU/2/4/896B), Uganda, Gulu University Multifunctional Laboratories (GUREC-093-20), Makerere University, College of Health Science, Kamala, Uganda (SBS-2022-130) and Zambia, Tropical Diseases Research Centre, Ndola Teaching Hospital (00003729). In all other countries, IRB approval for re-testing anonymized specimens was not required. Scripts for data analyses and data are available at GitHub (https://github.com/CarloFischer88/Analyse-the-BA.1-spread-in-Africa.git).
Phylogenetic analyses of African BA.1 sequences (Fig. S6) and phylogeographic analyses (Fig. 3, Fig. S7) are based on 942 sequences available on GISAID, via 10.55876/gis8.230818aq.
The performance of the used real-time RT-PCR assays was validated in this study against SARS-CoV-2 genomes generated by HTS (https://github.com/CarloFischer88/Analyse-the-BA.1-spread-in-Africa.git). Those genomes that were submitted to GISAID (Botswana, Guinea, South Africa) are available via 10.55876/gis8.240201eb. Mapped genomic reads for SARS-CoV-2 genomes generated from Beninese samples are available via the European Nucleotide Archive (ENA) (Project accession number PRJEB64297).

https://github.com/CarloFischer88/Analyse-the-BA.1-spread-in-Africa.git

## Acknowledgments

We thank our colleagues who provided insight and expertise that greatly assisted the research:

Sarah Belkalem, National Influenza Centre, Viral Respiratory Laboratory, Department of Virology, Institut Pasteur of Algeria, Algeria

El Alia Gradi, National Influenza Centre, Viral Respiratory Laboratory, Department of Virology, Institut Pasteur of Algeri, Algeria

Kahina Izri, National Influenza Centre, Viral Respiratory Laboratory, Department of Virology, Institut Pasteur of Algeri, Algeria

Sonia Carvalho, Laboratório de Biologia Molecular, Instituto Nacional de Investigação em Saúde (INIS), Luanda, Angola

Joana P. da Paixão, Laboratório de Biologia Molecular, Instituto Nacional de Investigação em Saúde (INIS), Luanda, Angola

Susana Daniel, Laboratório de Biologia Molecular, Instituto Nacional de Investigação em Saúde (INIS), Luanda, Angola

Kumbelembe David, Laboratório de Biologia Molecular, Instituto Nacional de Investigação em Saúde (INIS), Luanda, Angola

Moisés Dembo, Laboratório de Biologia Molecular, Instituto Nacional de Investigação em Saúde (INIS), Luanda, Angola

Julio Estobre, Laboratório de Biologia Molecular, Instituto Nacional de Investigação em Saúde (INIS), Luanda, Angola

Luzia Inglês, Laboratório de Biologia Molecular, Instituto Nacional de Investigação em Saúde (INIS), Luanda, Angola

Domingos Jandondo, Laboratório de Biologia Molecular, Instituto Nacional de Investigação em Saúde (INIS), Luanda, Angola

Agostinho Paulo, Laboratório de Biologia Molecular, Instituto Nacional de Investigação em Saúde (INIS), Luanda, Angola

Amilton Pereira, Laboratório de Biologia Molecular, Instituto Nacional de Investigação em Saúde (INIS), Luanda, Angola

Ramaliatou Chabi Nari, Laboratoire des fievres hemorragiques virales de Cotonou; Akpakpa, Cotonou, Benin

Rejeunie P.J. Mindzie Ngomo, Laboratoire des fievres hemorragiques virales de Cotonou; Akpakpa, Cotonou, Benin

Stéphane Sohou, Laboratoire des fievres hemorragiques virales de Cotonou; Akpakpa, Cotonou, Benin

Ragive Parode Takale, Molecular diagnostic Laboratory HDL, Pointe-Noire, Congo Negeri Debela, Hirsch Institute of Tropical Medicine, Asella, Ethiopia

Friederike Hunstig, Department of Gastroenterology, Hepatology and Infectious Diseases, University Hospital Düsseldorf, Heinrich Heine University Düsseldorf, Germany & Hirsch Institute of Tropical Medicine, Asella, Ethiopia

Tom Lüdde, Department of Gastroenterology, Hepatology and Infectious Diseases, University Hospital Düsseldorf, Heinrich Heine University Düsseldorf, Germany & Hirsch Institute of Tropical Medicine, Asella, Ethiopia

Julia Cyrielle Andeko, Centre Interdisciplinaire de Recherches Médicales de Franceville (CIRMF), Gabon

Lucie Marquet, Centre Interdisciplinaire de Recherches Médicales de Franceville (CIRMF), Gabon

Mary Basiru Njai, Medical Research Council Unit at London School of Hygiene and Tropical Medicine, Gambia

Ebrima Ceesay, Medical Research Council Unit at London School of Hygiene and Tropical Medicine, Gambia

Fatoumatta Cham, Medical Research Council Unit at London School of Hygiene and Tropical Medicine, Gambia

Hoja Gaye, Medical Research Council Unit at London School of Hygiene and Tropical Medicine, Gambia

Yusupha Jallow, Medical Research Council Unit at London School of Hygiene and Tropical Medicine, Gambia

Mamlie Touray, Medical Research Council Unit at London School of Hygiene and Tropical Medicine, Gambia

Mariama Touray, Medical Research Council Unit at London School of Hygiene and Tropical Medicine, Gambia

Wendy Karen Jó Lei, Charité – Universitätsmedizin Berlin, corporate member of Freie Universität Berlin and Humboldt Universität zu Berlin, Institute of Virology, Germany

Andrés Enrique Moreira-Soto, Charité – Universitätsmedizin Berlin, corporate member of Freie Universität Berlin and Humboldt Universität zu Berlin, Institute of Virology, Germany

Anna-Lena Sander, Charité – Universitätsmedizin Berlin, corporate member of Freie Universität Berlin and Humboldt Universität zu Berlin, Institute of Virology, Germany

Matthieu Domenech de Cellès, Infectious Disease Epidemiology Group, Max Planck Institute for Infection Biology, Berlin, Germany

Henry Acheampong, KCCR, UPO, PMB, KNUST, Kumasi, Ghana

Millicent Afatodzie, Noguchi Memorial Institute for Medical Research, Ghana

Sherihane Aryeetey, KCCR, UPO, PMB, KNUST, Kumasi, Ghana

Christopher Dorcoo, Noguchi Memorial Institute for Medical Research, University of Ghana, Ghana

Elvis Lomotey, Noguchi Memorial Institute for Medical Research, University of Ghana, Ghana

Daniel Odumang, Noguchi Memorial Institute for Medical Research, University of Ghana, Ghana

Millicent Opoku, Noguchi Memorial Institute for Medical Research, University of Ghana, Ghana

Grace Opoku-Gyamfi, Noguchi Memorial Institute for Medical Research, University of Ghana, Ghana

Millicent Oye Kyei, Noguchi Memorial Institute for Medical Research, University of Ghana, Ghana

Shalyn Akasa, National Public Health Reference Laboratory, Ministry of Health, Kenya

Claudio Raharinandrasana, Institu Pasteur de Madagascar - Virology Unit, Madagaskar

Anne-Marie Ratsimbazafy, Institu Pasteur de Madagascar - Virology Unit, Madagaskar

Idrissa Coulibaly, Health Clinic, Sanso, Morila SA, Mali

Mamoudou Kanoute, Health Clinic, SOMISY, Mali

Mama Kanta, Health Clinic, SOMILO, Mali

Aminata Maiga, Laboratoire d’analyses médicales, CHU Point G, Bamako, Mali, Mali

Didier Ndane, Health Clinic, SMK, Mali, Mali

Mamadou Sanghata, Health Clinic, Sanso, Morila SA, Mali

Abdoul Aziz Sow, Health Clinic, SOMILO, Mali

Abdelmajid Eloualid, Institut Pasteur du Maroc, Casablanca, Marocco

Abdellah Faouzi, Institut Pasteur du Maroc, Casablanca, Marocco

Saloua Nadifiyine, Institut Pasteur du Maroc, Casablanca, Marocco

Deborah Goudiaby, Institut Pasteur de Dakar (IPD), Senegal

Davy Evrard Kiori, Institut Pasteur de Dakar (IPD), Senegal

Marie Pedapa Mendy, Institut Pasteur de Dakar (IPD), Senegal

Solène Grayo, Institut Pasteur de Guinée (IPGui), Guinée

Yann Le Pennec, Institut Pasteur de Guinée (IPGui), Guinée

Reine Salomé Anguinze, Institut Pasteur de Guinée (IPGui), Guinée

Mathilda Claassen, Stellenbosch University, South Africa

Bronwyn Roberts, Stellenbosch University, South Africa

Shannon Wilson, Stellenbosch University and National Health Laboratory Service Tygerberg, South Africa

Fortune Salah, Institut National d’Hygiène, Lomé, Togo

Micheline Tettekpoe, Institut National d’Hygiène, Lomé, Togo

Frida Aryemo, Gulu University Multifunctional Laboratories, Gulu, Uganda

Doreen Ato, Gulu University Multifunctional Laboratories, Gulu, Uganda

Ian Goodhead, Gulu University Multifunctional Laboratories, Gulu, Uganda and School of Science, Engineering and EnvironmPrunusent, University of Salford, United Kingdom

Pamela Lamwaka, Gulu University Multifunctional Laboratories, Gulu, Uganda

We gratefully acknowledge all data contributors, i.e., the authors and their originating laboratories responsible for obtaining the specimens, and their submitting laboratories for generating the genetic sequence and metadata and sharing via the GISAID Initiative, on which parts of this research are based.

## Funding

African Academy of Sciences grant SARSCov2-4-20-004 (IOD)

AFROSCREEN project (grant agreement CZZ3209) through Agence Française de Développement, coordinated by ANRS | Maladies infectieuses émergentes in partnership with Institut Pasteur and IRD (IPdG). We would additionally like to thank members from the AFROSCREEN Consortium (https://www.afroscreen.org/en/network/) for their work and support on genomic surveillance in Africa.

Bill & Melinda Gates Foundation grant INV-005971 (JFD, CD); grant INV-024130 (IOD); The findings and conclusions contained within are those of the authors and do not necessarily reflect positions or policies of the Bill & Melinda Gates Foundation CIRMF is a member of CANTAM funding by EDCTP CSA2020NoE-3100 – CANTAM 3 and supported by the Gabonese Government and Total Gabon

French Ministry of Europe and Foreign affairs (Bamako-Bordeaux decentralized cooperation in riposte to Covid-19)

Horizon 2020, European and Developing Countries Clinical Trials Partnership (EDCTP2) programme, PANDORA-ID-NET Grant RIA2016E-1609 (AASy, ROP)

Poliomyelitis Research Foundation grant 21/40 (KKP)Programa de Desenvolvimento de Ciência e Tecnologia grant N°11/MESCTI/PDCT/2020 (JFMdM)

“REPAIR Covid-19-Africa,” coordinated by the Pasteur Network association and funded by French Ministry for Europe and Foreign Affairs (MEAE), funded COVID diagnosis and typing (IPdG).

South African Department of Science and Innovation (sub-award via University of KwaZulu-Natal) grant S006872 (TGM)

Stellenbosch University Postgraduate Scholarship Programme grant 25095676 (KKP) UKRI Global Challenges Research Fund, grant number NF118 (RB, RE, JMw) World Health Organization grant 2021/1113013-0 (IOD)

## Author contributions

Resources: TGM, AY, NA, EA, PA, PAf, JA, SFA, LA, YA, MAB, AB, RB, ALMB, FB, MC, PC, RMC, JC, GC, AC, UDA, XNdL, JFMdM, FD, ND, YD, LD, PD, RE, YE, AE, OF, TF, AH, PVI, NI, RJ, SJ, BK, JK, LK, OK, VL, AL, OL, SELD, JBLD, EL, HL, JL, SM, IM, BM, PAM, JM, LM, JMw, NN, CAN, MON, EM, RN, JN, SGN, EOO, AO, JBO, MO IOD, KKP, ROP, WP, VR, FS, SS, AAS, AASy, PATN, ZT, FOT, TBT, PR, NT, SMo, FC, WC.

Supervision: JFD

Validation: CF, TGM, AY, JFD

Visualization: CF

Writing – original draft: CF, JFD

Writing – review & editing: CF, TGM, AY, JFD

Conceptualization: CF, JFD

Data curation: CF, AF

Formal analysis: CF, AF, JFD

Funding acquisition: TGM, RB, JFMdM, CD, IOD, KKP, ROP, AASy, JFD

Investigation CF, TGM, AY, NA, EA, PA, PAf, JA, SFA, LA, YA, MAB, AB, RB, ALMB, FB, MC, PC, RMC, JC, GC, AC, UDA, XNdL, JFMdM, FD, ND, YD, LD, PD, RE, YE, AE, OF, TF, AH, PVI, NI, RJ, SJ, BK, JK, LK, OK, VL, AL, OL, SELD, JBLD, EL, HL, JL, SM, IM, BM, PAM, JM, LM, JMw, NN, CAN, MON, EM, RN, JN, SGN, EOO, AO, JBO, MO IOD, KKP, ROP, WP, VR, FS, SS, AAS, AASy, PATN, ZT, FOT, TBT, FC, PR, NT, WC, SMo, SG.

Methodology: CF, JFD

Project administration: AK, JFD

## Competing interests

Olfert Landt is the former owner of TIB Molbiol, the company that produced the kits provided to African partner laboratories within the framework of this study. The kits are not commercially available. All authors declare that they have no competing interests.

## Data and materials availability

Respiratory samples sent to laboratories for COVID-19 testing by attending physicians were re-tested in an anonymized fashion using the COVID-19 test developed in this study. Ethical approval for re-testing and scientific usage was provided by the institutional research ethics board (IRB) of Charité-Universitätsmedizin Berlin (EA2/028/22) and by IRBs from Burkina Faso, Laboratoire National de Référence-Grippes (2020-7-126), Cameroon, Centre Pasteur du Cameroun (2020/05/1224/CE/CNERSH/SP), Ghana, Kumasi Centre for Collaborative Research in Tropical Medicine (KCCR), KNUST (CHRPE/AP/566/21), Kenya, Jomo Kenyatta University of Agriculture and Technology, Department of Biochemistry (JKU/2/4/896B), Uganda, Gulu University Multifunctional Laboratories (GUREC-093-20), Makerere University, College of Health Science, Kamala, Uganda (SBS-2022-130) and Zambia, Tropical Diseases Research Centre, Ndola Teaching Hospital (00003729). In all other countries, IRB approval for re-testing anonymized specimens was not required. Scripts for data analyses and data are available at GitHub (https://github.com/CarloFischer88/Analyse-the-BA.1-spread-in-Africa.git).

Phylogenetic analyses of African BA.1 sequences (**Fig. S6**) and phylogeographic analyses (**Fig. 3**, **Fig. S7**) are based on 942 sequences available on GISAID, via 10.55876/gis8.230818aq.

The performance of the used real-time RT-PCR assays was validated in this study against SARS-CoV-2 genomes generated by HTS (https://github.com/CarloFischer88/Analyse-the-BA.1-spread-in-Africa.git). Those genomes that were submitted to GISAID (Botswana, Guinea, South Africa) are available via 10.55876/gis8.240201eb. Mapped genomic reads for SARS-CoV-2 genomes generated from Beninese samples are available via the European Nucleotide Archive (ENA) (Project accession number PRJEB64297).

## Notes

### Competing Interest Statement

The authors have declared no competing interest.

### Funding Statement

African Academy of Sciences grant SARSCov2-4-20-004 (IOD)
AFROSCREEN project (grant agreement CZZ3209) through Agence Francaise de Developpement, coordinated by ANRS | Maladies infectieuses emergentes in partnership with Institut Pasteur and IRD (IPdG). We would additionally like to thank members from the AFROSCREEN Consortium (https://www.afroscreen.org/en/network/) for their work and support on genomic surveillance in Africa.
Bill & Melinda Gates Foundation grant INV-005971 (JFD, CD); grant INV-024130 (IOD); The findings and conclusions contained within are those of the authors and do not necessarily reflect positions or policies of the Bill & Melinda Gates Foundation
CIRMF is a member of CANTAM funding by EDCTP CSA2020NoE-3100 - CANTAM 3 and supported by the Gabonese Government and Total Gabon
French Ministry of Europe and Foreign affairs (Bamako-Bordeaux decentralized cooperation in riposte to Covid-19)
Horizon 2020, European and Developing Countries Clinical Trials Partnership (EDCTP2) programme, PANDORA-ID-NET Grant RIA2016E-1609 (AASy, ROP)
Poliomyelitis Research Foundation grant 21/40 (KKP)Programa de Desenvolvimento de Ciencia e Tecnologia grant No11/MESCTI/PDCT/2020 (JFMdM)
REPAIR Covid-19-Africa, coordinated by the Pasteur Network association and funded by French Ministry for Europe and Foreign Affairs (MEAE), funded COVID diagnosis and typing (IPdG).
South African Department of Science and Innovation (sub-award via University of KwaZulu-Natal) grant S006872 (TGM)
Stellenbosch University Postgraduate Scholarship Programme grant 25095676 (KKP)
UKRI Global Challenges Research Fund, grant number NF118 (RB, RE, JMw)
World Health Organization grant 2021/1113013-0 (IOD)

### Author Declarations

Ethical approval for re-testing and scientific usage was provided by the institutional research ethics board (IRB) of Charite-Universitaetsmedizin Berlin (EA2/028/22) and by IRBs from Burkina Faso, Laboratoire National de Reference-Grippes (2020-7-126), Cameroon, Centre Pasteur du Cameroun (2020/05/1224/CE/CNERSH/SP), Ghana, Kumasi Centre for Collaborative Research in Tropical Medicine (KCCR), KNUST (CHRPE/AP/566/21), Kenya, Jomo Kenyatta University of Agriculture and Technology, Department of Biochemistry (JKU/2/4/896B), Uganda, Gulu University Multifunctional Laboratories (GUREC-093-20), Makerere University, College of Health Science, Kamala, Uganda (SBS-2022-130) and Zambia, Tropical Diseases Research Centre, Ndola Teaching Hospital (00003729).

