## Supplementary Data for "Ineffectiveness of international travel restrictions to contain spread of the SARS-CoV-2 Omicron BA.1 variant: a continent-wide laboratory-based observational study from Africa"

### Supplementary tables

#### Table S1.

| Oligonucleotide | Sequence | Target |
|---|---|---|
| E-Sarbecco-F1 | ACAggTACgTTAATAgTTAATAgCgT | SARS-CoV-2 |
| E-Sarbecco-F2 | gCTTTCgTggTATTCTTgCTAg | |
| E-Sarbecco-R1 | CAATATTgCAgCAgTACgCACA | |
| E-Sarbecco-P | 610-ACACTAgCCATCCTTACTgCgCTTCg—Q | |
| del157/158-F | CAATTTTgTAATgATCCATTTTTggg | Delta variant |
| del157/158-R | ATATTCAAAAgTgCAATTATTCgC | |
| del157/158-P | 580-TagAA+TAAACTCCACTTTCCA+TCC—Q | |
| ins214EPE-S | AAATATATTCTAAgCACACgCCT | BA.1 variant |
| ins214EPE-B | gTTCTAAAgCCgAAAAACCCTg | |
| ins214EPE-P | FAM-AgTgCgTgAgCCAgAAgATCTCC—Q | |

Real-time RT-PCR assays provided to participants. The recommended cycling protocol included reverse transcription for 3 minutes at 55°C, initial denaturation for 1 minute at 95°C, 40 cycles of 95°C for 3 seconds and 63°C for 10 seconds and cool down at 40°C for 10 seconds. Locked nucleic acids are indicated by + symbols. PCR kits with lyophilized enzymes to ensure robust testing under tropical conditions were made available during the BA.1 wave in December 2021 free of charge and unconditionally. The PCR kits were designed specifically for this study and are not commercially available.

#### Table S2.

| Variant | Genetic marker | N sequences with marker | N variant | N variant with marker | Analytical sensitivity | Analytical specificity |
|---|---|---|---|---|---|---|
| Delta | S:Del157/158 | 3,859,121 | 3,689,153 | 3,648,794 | 94.5% | 98.9% |
| BA.1 | S:Ins214EPE | 295,713 | 375,536 | 291,143 | 77.5% | 98.5% |

Performance analyses of the provided typing PCR kits by all available SARS-CoV-2 sequences available on GISAID on January 18, 2022. The specificity of the selected markers was 98.9% for typing of the Delta variant and 98.5% for typing of the BA.1 variant based on all GIASAID sequences available on January 18, 2022. Sequences containing >5% of Ns were excluded to ensure high sequence quality among GISAID entries.

**Table S3.**

| Country | Mean age | Female | Male | Sex unknown | Sampling sites |
|---|---|---|---|---|---|
| Algeria | 51.8 | 50·1 | 49·9 | 0·0 | 11 |
| Angola | 35.1 | 40·0 | 60·0 | 0·0 | 1 |
| Benin | 37.9 | 39·6 | 57·7 | 2·7 | 1 |
| Botswana | 30.4 | 36·4 | 25·1 | 38·5 | 23 |
| Burkina Faso | 39.5 | 29·9 | 70·1 | 0·0 | 2 |
| Cameroon | 40.2 | 50.1 | 49.2 | 0.7 | 1 |
| Ethiopia | 40.5 | 32.8 | 63.4 | 3.8 | 16 |
| Gabon | 40.8 | 43.0 | 57.0 | 0.0 | 2 |
| Gambia | 35.3 | 47.1 | 52.9 | 0.0 | 1 |
| Ghana | 33.8 | 44.2 | 51.8 | 4.0 | 4 |
| Guinea | 35.6 | 48.4 | 51.1 | 0.5 | 5 |
| Kenya | 41.0 | 45.3 | 54.7 | 0.0 | 9 |
| Madagascar | 36.9 | 55.7 | 43.3 | 1.0 | 27 |
| Mali | 37.8 | 13.4 | 74.3 | 12.3 | 5 |
| Morocco | 44.5 | 53.6 | 45.8 | 0.6 | 2 |
| Mozambique | 33.4 | 56.5 | 43.5 | 0.0 | 24 |
| Namibia | 38.5 | 47.0 | 53.0 | 0.0 | 4 |
| Niger | 39.6 | 27.8 | 66.7 | 5.5 | 3 |
| Republic of the Congo | 38.1 | 16.8 | 83.2 | 0.0 | 1 |
| Senegal | 38.1 | 47.2 | 52.8 | 0.0 | 9 |
| South Africa | 39.1 | 53.6 | 43.7 | 2.7 | 5 |
| Togo | 38.5 | 39.0 | 60.7 | 0.3 | 20 |
| Uganda | 34.0 | 47.1 | 52.4 | 0.5 | 48 |
| Zimbabwe | 42.3 | 53.7 | 46.3 | 0.0 | 1 |

Description of the study cohort.

**Table S4**.

| | glm1 | glm2 | glm3 | glm4 | glm5 | glm6 | glm7 |
|---|---|---|---|---|---|---|---|
| Applied formula | y ~ x | y ~ x^2 | y ~ x + x^2 | y ~ x^3 | y ~ x + x^3 | y ~ x/2 | y ~ x*2 |
| Algeria | 1.0000 | 1.0000 | 1.0060 | 1.0016 | 1.0060 | 1.0000 | 1.0000 |
| Angola | 1.3794 | 1.4525 | 1.0000 | 1.5325 | 1.0040 | 1.3794 | 1.3794 |
| Benin | 1.2162 | 1.1167 | 1.0364 | 1.0348 | 1.0000 | 1.2162 | 1.2162 |
| Botswana | 1.0926 | 1.1107 | 1.0000 | 1.1327 | 1.0023 | 1.0926 | 1.0926 |
| Burkina Faso | 1.0000 | 1.0021 | 1.0539 | 1.0046 | 1.0540 | 1.0000 | 1.0000 |
| Cameroon | 1.1026 | 1.1313 | 1.0000 | 1.1692 | 1.0036 | 1.1026 | 1.1026 |
| Ethiopia | 1.0000 | 1.0000 | 1.1325 | 1.0000 | 1.1325 | 1.0000 | 1.0000 |
| Gambia | 1.5486 | 1.6480 | 1.0000 | 1.7653 | 1.0059 | 1.5486 | 1.5486 |
| Ghana | 1.0352 | 1.0392 | 1.0000 | 1.0440 | 1.0015 | 1.0352 | 1.0352 |
| Guinea | 1.0000 | 1.0241 | 3.4543 | 1.0502 | 3.4543 | 1.0000 | 1.0000 |
| Kenya | 1.0000 | 1.0069 | 1.0414 | 1.0142 | 1.0433 | 1.0000 | 1.0000 |
| Madagascar | 1.1667 | 1.1882 | 1.0000 | 1.2119 | 1.0009 | 1.1667 | 1.1667 |
| Mali | 1.0286 | 1.0152 | 1.0000 | 1.0075 | 1.0025 | 1.0286 | 1.0286 |
| Morocco | 1.0000 | 1.0005 | 1.0047 | 1.0012 | 1.0047 | 1.0000 | 1.0000 |
| Mozambique | 1.0000 | 1.0050 | 1.0070 | 1.0133 | 1.0073 | 1.0000 | 1.0000 |
| Namibia | 1.6280 | 1.7196 | 1.0000 | 1.8244 | 1.0054 | 1.6280 | 1.6280 |
| Niger | 1.0035 | 1.0071 | 1.0002 | 1.0116 | 1.0000 | 1.0035 | 1.0035 |
| Republic of the Congo | 1.0945 | 1.1229 | 1.0000 | 1.1566 | 1.0010 | 1.0945 | 1.0945 |
| Senegal | 1.0056 | 1.0024 | 1.0067 | 1.0000 | 1.0057 | 1.0056 | 1.0056 |
| South Africa | 1.2857 | 1.3731 | 1.0000 | 1.4843 | 1.0174 | 1.2857 | 1.2857 |
| Togo | 1.0000 | 1.0070 | 1.0017 | 1.0177 | 1.0018 | 1.0000 | 1.0000 |
| Uganda | 1.0124 | 1.0190 | 1.0015 | 1.0291 | 1.0000 | 1.0124 | 1.0124 |
| **Summed AICs** | 24.5996 | 24.9915 | 24.7463 | 25.5067 | 24.7541 | 24.5996 | 24.5996 |
| **Mean AIC** | 1.1182 | 1.1360 | 1.1248 | 1.1594 | 1.1252 | 1.1182 | 1.1182 |
| **Median AIC** | 1.0205 | 1.0215 | 1.0009 | 1.0319 | 1.0044 | 1.0205 | 1.0205 |

Performance of generalized linear models to predict the PCR-based BA.1 fraction over time by Akaike information criterion (AIC).

### Table S5.

| Simu-lation | Johannesburg | | | Cape Town | | | Durban | | | Gabore | | | Total cases | Factor |
|---|---|---|---|---|---|---|---|---|---|---|---|---|---|---|
| | mild | severe | Total | mild | severe | Total | mild | severe | Total | mild | severe | Total | | |
| 1 | 337 | 37 | 374 | 90 | 10 | 100 | 90 | 10 | 100 | 320 | 36 | 355 | 929 | 1.00 |
| 2 | 505 | 56 | 561 | 135 | 15 | 150 | 135 | 15 | 150 | 479 | 53 | 533 | 1,394 | 1.50 |
| 3 | 757 | 84 | 842 | 203 | 23 | 225 | 203 | 23 | 225 | 719 | 80 | 799 | 2,090 | 2.25 |
| 4 | 1,136 | 126 | 1,262 | 304 | 34 | 338 | 304 | 34 | 338 | 1,078 | 120 | 1,198 | 3,135 | 3.38 |
| 5 | 1,704 | 189 | 1,893 | 456 | 51 | 506 | 456 | 51 | 506 | 1,617 | 180 | 1,797 | 4,703 | 5.06 |
| 6 | 2,556 | 284 | 2,840 | 683 | 76 | 759 | 683 | 76 | 759 | 2,426 | 270 | 2,696 | 7,055 | 7.59 |
| 7 | 3,834 | 426 | 4,260 | 1,025 | 114 | 1,139 | 1,025 | 114 | 1,139 | 3,639 | 404 | 4,044 | 10,582 | 11.39 |
| 8 | 5,751 | 639 | 6,390 | 1,538 | 171 | 1,709 | 1,538 | 171 | 1,709 | 5,459 | 607 | 6,066 | 15,873 | 17.09 |
| 9 | 8,627 | 959 | 9,585 | 2,307 | 256 | 2,563 | 2,307 | 256 | 2,563 | 8,188 | 910 | 9,098 | 23,809 | 25.63 |
| 10 | 12,940 | 1,438 | 14,378 | 3,460 | 384 | 3,844 | 3,460 | 384 | 3,844 | 12,283 | 1,365 | 13,647 | 35,714 | 38.44 |
| 11 | 19,410 | 2,157 | 21,566 | 5,190 | 577 | 5,766 | 5,190 | 577 | 5,766 | 18,424 | 2,047 | 20,471 | 53,571 | 57.67 |
| 12 | 29,115 | 3,235 | 32,350 | 7,785 | 865 | 8,649 | 7,785 | 865 | 8,649 | 27,636 | 3,071 | 30,707 | 80,356 | 86.50 |

Defined starting BA.1 cases in GLEAMviz simulations.

### Table S6.

| | HTS result | | | Clinical PCR performance | |
|---|---|---|---|---|---|
| PCR result | Delta | Omicron/BA.1 | Other | Sensitivity | Specificity |
| Delta | 293 | 0 | 0 | 92.4% | 100% |
| Omicron/BA.1 | 3 | 475 | 5 | 99.6% | 97.6% |
| Other | 20 | 2 | 12 | | |
| Negative | 1 | 0 | 0 | | |

Validation of the typing PCR assays with clinical samples from Benin, Botswana, Guinea, and South Africa.

### *Table S7.*

| Region | Filtered data | Raw filtered data |
|---|---|---|
| Southern Africa | 08.11.2021 (05.11.2021 – 11.11.2021) | 08.11.2021 (05.11.2021 – 11.11.2021) |
| Eastern Africa | 13.12.2021 (10.12.2021 – 16.12.2021) | 12.12.2021 (09.12.2021 – 15.12.2021) |
| Central Africa | 10.12.2021 (07.12.2021 – 13.12.2021) | 11.12.2021 (09.12.2021 – 14.12.2021) |
| Northern Africa | 25.12.2021 (24.12.2021 – 26.12.2021) | 25.12.2021 (24.12.2021 – 26.12.2021) |
| Western Africa | 01.12.2021 (30.11.2021 – 02.12.2021) | 29.11.2021 (27.11.2021– 30.11.2021) |

Modelled date of BA.1 dominance based on raw and filtered PCR results.

**Table S8.**

| Country | First BA.1 positive sample by qPCR | First expected BA.1 positive sample from date of BA.1 dominance |
|---|---|---|
| Algeria | 30.11.2021 | 30.10.2021 |
| Angola | 17.12.2021 | 19.08.2021 |
| Benin | 03.08.2021 | 03.09.2021 |
| Botswana | 12.11.2021 | 21.09.2021 |
| Burkina Faso | 13.12.2021 | 14.10.2021 |
| Cameroon | 01.12.2021 | 02.10.2021 |
| Ethiopia | 21.11.2021 | 16.10.2021 |
| Gabon | - | - |
| Gambia | 14.12.2021 | 20.08.2021 |
| Ghana | 02.09.2021 | 20.09.2021 |
| Guinea | 20.12.2021 | 08.10.2021 |
| Kenya | 20.12.2021 | 12.10.2021 |
| Madagascar | 10.01.2022 | 11.11.2021 |
| Mali | 02.08.2021 | 02.10.2021 |
| Morocco | 15.12.2021 | 11.10.2021 |
| Mozambique | 19.11.2021 | 11.09.2021 |
| Namibia | 29.11.2021 | 12.09.2021 |
| Niger | 03.09.2021 | 26.10.2021 |
| Republic of the Congo | 17.12.2021 | 07.10.2021 |
| Senegal | 23.11.2021 | 04.10.2021 |
| South Africa | 26.11.2021 | 29.08.2021 |
| Togo | 21.11.2021 | 02.10.2021 |
| Uganda | 01.09.2021 | 30.09.2021 |
| Zimbabwe | 24.11.2021 | 14.10.2021 |

Collection dates of first BA.1 positive samples and dates of first expected BA.1 cases based on the timepoint of BA.1 becoming the dominant variant, population sizes and an assumed doubling time of 3 days.

**Table S9**.

| Country | Median Rt |
|---|---|
| Algeria | 1.80 (1.64 - 1.98) |
| Benin | 2.46 (2.21 - 2.67) |
| Burkina Faso | 2.99 (2.67 - 3.19) |
| Cameroon | 1.24 (1.23 - 1.29) |
| Republic of Congo | 1.10 (1.08 - 1.13) |
| Ethiopia | 3.67 (3.64 - 3.70) |
| Ghana | 2.40 (2.26 - 2.56) |
| Guinea | 2.98 (2.75 - 3.15) |
| Kenya | 2.72 (2.54 - 2.88) |
| Madagascar | 2.21 (1.90 - 2.56) |
| Mali | 1.33 (1.30 - 1.38) |
| Morocco | 2.92 (2.66 - 3.10) |
| Mozambique | 1.87 (1.74 - 1.99) |
| South Africa | 1.38 (1.34 - 1.44) |
| Senegal | 3.32 (3.06 - 3.46) |
| Togo | 2.46 (2.21 - 2.67) |
| Uganda | 2.12 (2.01 - 2.29) |

Median $R_t$ during the 30 days before BA.1 became the dominant SARS-CoV-2 variant in African countries considered for the $R_t$ analyses in Figure 4B.

**Table S10.**

| Country | Collection date | Omicron/BA.1 | Departure country |
|---|---|---|---|
| Togo | 11.2021 | 1 | Burkina Faso |
| Togo | 11.2021 | 1 | Nigeria |
| Senegal | 11.2021 | 1 | Mauritania |
| Togo | 11.2021 | 1 | Niger |
| Algeria | 11.2021 | 1 | South Africa |
| Togo | 12.2021 | 1 | Nigeria |
| Senegal | 12.2021 | 1 | South Africa |
| Senegal | 12.2021 | 1 | |
| Togo | 12.2021 | 1 | Germany |
| Senegal | 12.2021 | 1 | |
| Togo | 12.2021 | 1 | United states of America |
| Togo | 12.2021 | 1 | Democratic Republic of Congo |
| Togo | 12.2021 | 1 | Netherlands |
| Senegal | 12.2021 | 1 | |
| Togo | 12.2021 | 1 | Nigeria |
| Senegal | 12.2021 | 1 | Philippines |
| Senegal | 12.2021 | 1 | |
| Togo | 12.2021 | 1 | Nigeria |
| Togo | 12.2021 | 1 | Ethiopia |
| Togo | 12.2021 | 1 | Nauru |
| Senegal | 12.2021 | 1 | |
| Senegal | 12.2021 | 1 | |
| Senegal | 12.2021 | 1 | |
| Niger | 12.2021 | 1 | Australia |
| Togo | 12.2021 | 1 | Nauru |
| Togo | 12.2021 | 1 | Nauru |
| Togo | 12.2021 | 1 | Nauru |
| Togo | 12.2021 | 1 | France |
| Niger | 12.2021 | 1 | Côte d’Ivoire |

BA.1 detections in inbound travelers before December 10, 2021, when early BA.1 detections were summarized by the European Center for Disease Control and Prevention. Exact dates not shown to meet medRxiv data protection requirements.

***Table S11.***

| Region | PCR informed model | | Best fitting model | |
|---|---|---|---|---|
| | **Median Days (95% CI)** | **Date** | **Median Days (95% CI)** | **Date** |
| Southern Africa | 52 (26.0 – 58.0) | 02.01.2022 | 18.5 (8.0 – 21.0) | 29.11.2021 |
| Eastern Africa | 79 (69.0 – 87.0) | 29.01.2022 | 35 (27.5 - 50.5) | 16.12.2021 |
| Central Africa | 83 (73.0 – 101.0) | 02.02.2022 | 41 (24.0 – 68.0) | 22.12.2021 |
| Western Africa | 87 (74.0 – 100.0) | 06.02.2022 | 48 (37.0 – 65.0) | 29.12.2021 |
| Northern Africa | 92 (90.0 - 98.5) | 11.02.2022 | 52.5 (47.5 - 56.5) | 02.01.2022 |

First time points when one BA.1 infection in 100,000 inhabitants was simulated in the PCR informed model considering 13% pre-existing immunity against BA.1 and the best fitting model with 86.5-fold increased BA.1 starting cases.

***Table S12.***

| Approach | Reagent | Price ($) | Reactions | Price single reaction ($) | Total price per sample ($) | Hands-on time per sample | Advantages | Disadvantages |
|---|---|---|---|---|---|---|---|---|
| Real-time RT-PCR | LightCycler® Multiplex RNA Virus Master[a] | 2,701.07 | 1,000 | 2.70 | 7.82 | 1.9 minutes | Low costs, broadly applicable, short turnaround times, easy to upscale | Need for unique marker mutation, specific design for every variant needed, monitoring of marker specificity required, limited genomic information, performance may vary on different instruments |
| | Probe 1 (100 nmol)[b] | 205 | 2,000 | 0.10 | | | | |
| | Probe 2 (100 nmol)[b] | 205 | 2,000 | 0.10 | | | | |
| | Primers (2 sets)[b] | 32 | 2,000 | 0.02 | | | | |
| | MagNA Pure 96 DNA and Viral NA Small Volume Kit[c] | 2,821.46 | 576 | 4.90 | | | | |
| HTS | COVIDSeq Assay (96 samples) index 3 RUO[d] | 4,316.30 | 96 | 44.96 | 49.86 | 5.6 minutes | Complete genomic information, less variant specific | Not broadly available, long turnaround times, expensive, not easily upscaled, additional bioinformatical analyses required |
| | MagNA Pure 96 DNA and Viral NA Small Volume Kit[c] | 2,821.46 | 576 | 4.90 | | | | |

Example costs and time expenses for variant typing of one SARS-CoV-2 positive sample using real-time RT-PCR and HTS. Commonly used example products were chosen but cheaper and more expensive solutions are available for both approaches. Prices were requested and checked on June28, 2023. Hands-on times were calculated assuming a parallel test of 96 samples. References for the products:

[a] https://www.fishersci.com/shop/products/multiplex-rna-virus-master/NC1919522
[b] https://eu.idtdna.com/
[c] https://www.fishersci.com/shop/products/mp-96-dna-viral-na-sv-kt-ivd/50588381
[d] https://www.illumina.com/products/by-type/clinical-research-products/covidseq-assay.html

### Supplementary Figures

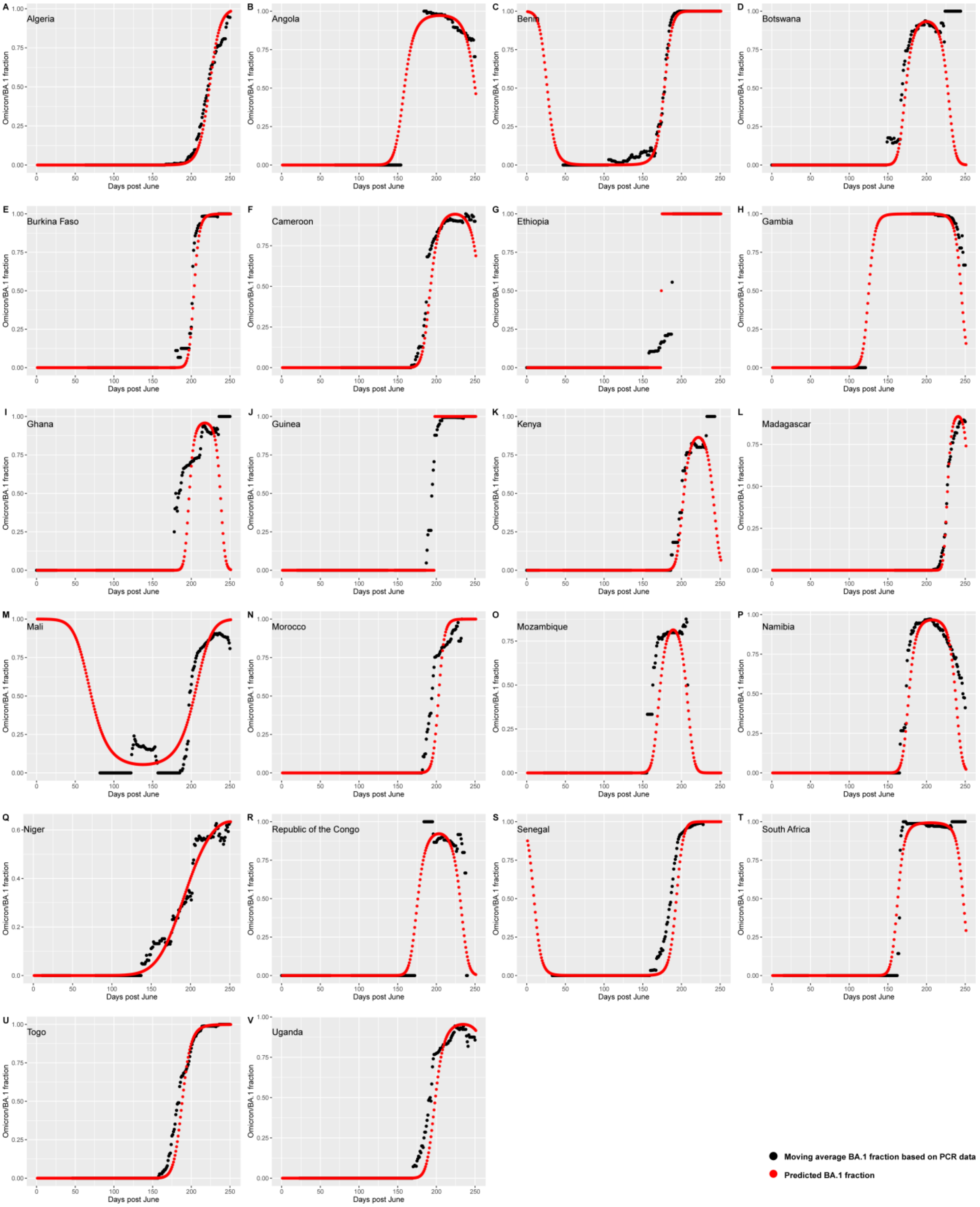


***Figure S1*: Prediction of the BA.1 fraction over time.** GLMs (red) were calculated using the formula "y ~ x + $x^2$". Moving average (black) was calculated with a window site of 21 days.

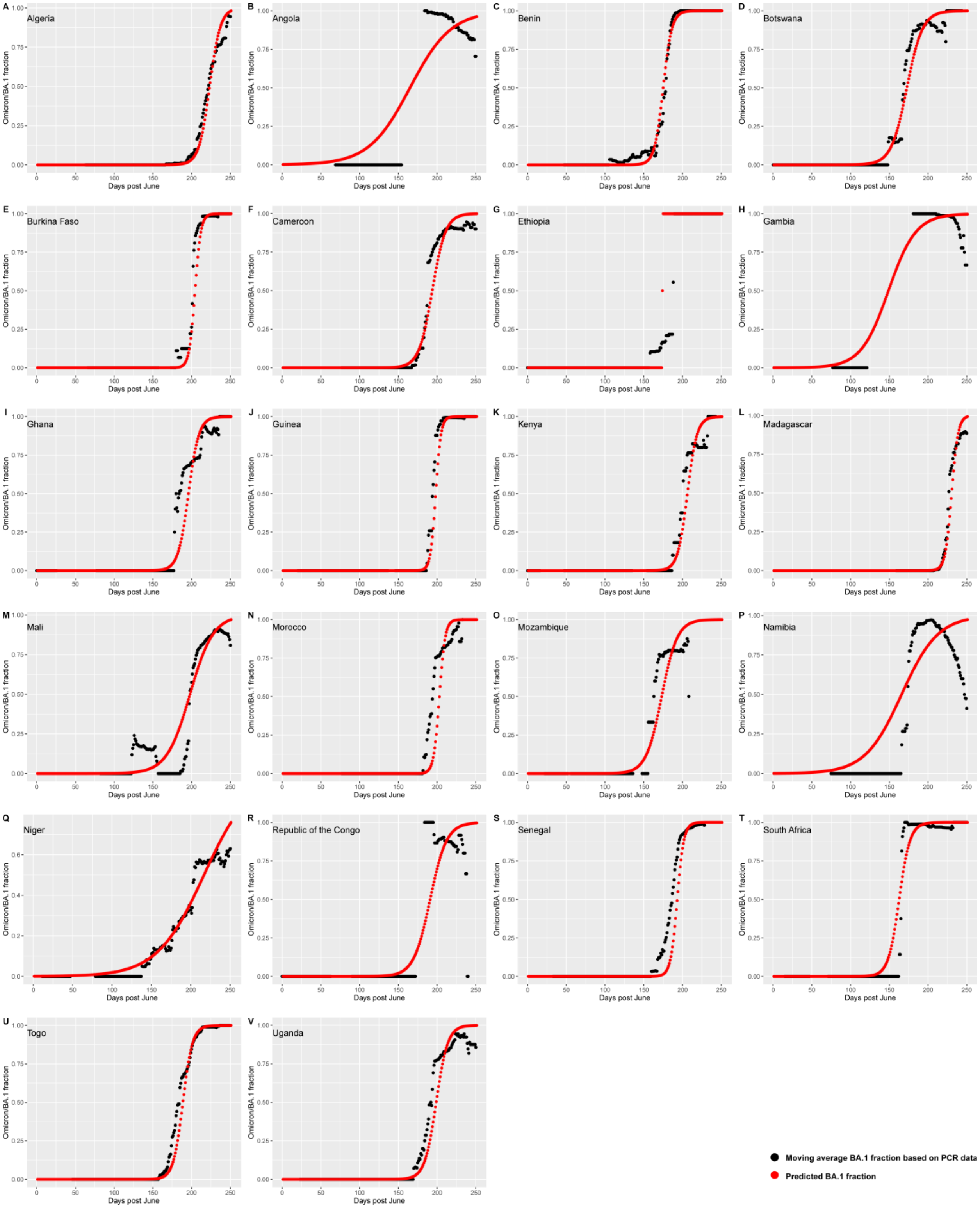


***Figure S2*: Prediction of the BA.1 fraction over time.** GLMs (red) were calculated using the formula “y ~ x". Moving average (black) was calculated with a window site of 21 days.

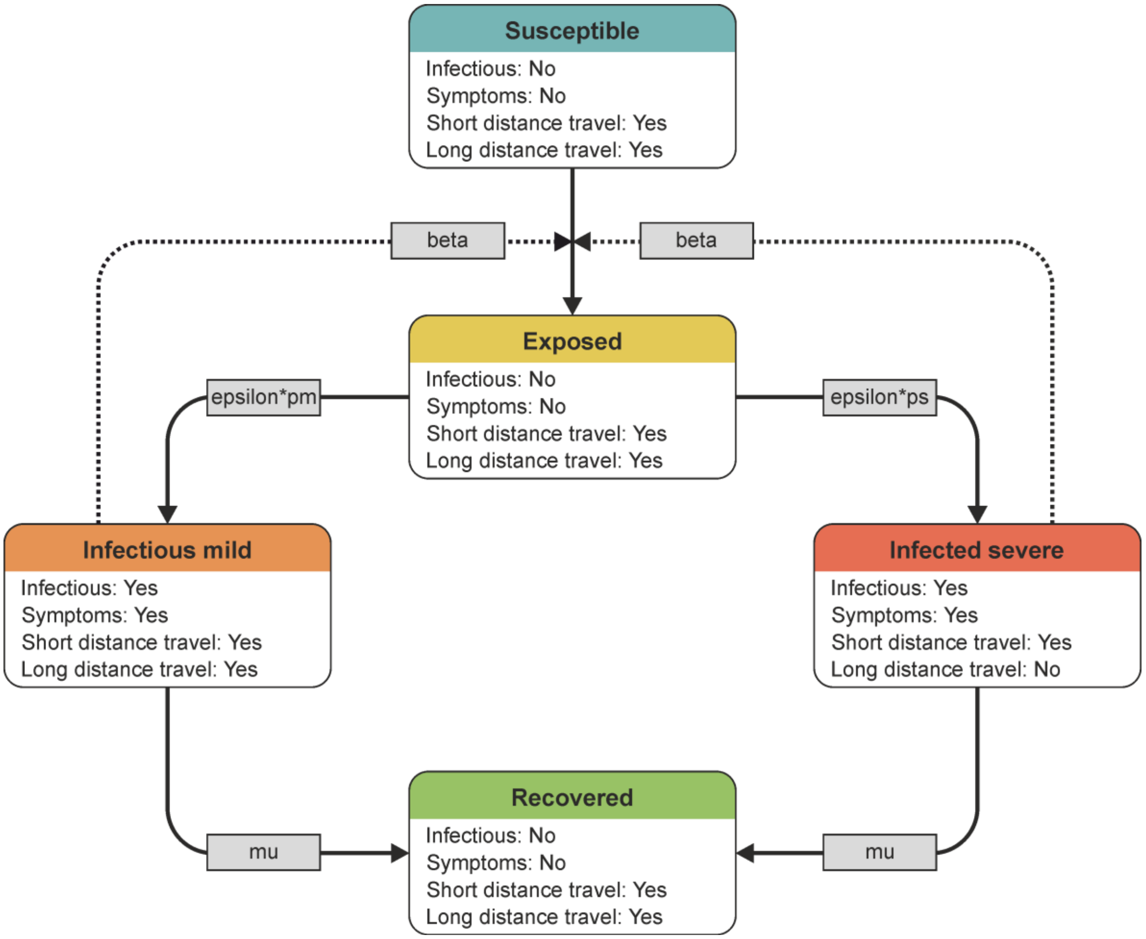


***Figure S3*: Setup of the "Susceptible", "Exposed", "Infected" and "Recovered" (SEIR) model in GLEAMviz.** Parameters were set as follows; Beta: Secondary attack rate (0.427)[33]. Epsilon: Transition from exposed and not infectious to infected (on average two days, 1/2)[34]. Pm: Fraction mild infections, air travel is not changed (90%). Ps: Fraction severe infections, severe symptoms lead to avoidance of air travel (10%). Mu: Duration of infectious virus shedding (on average 5 days, 1/5)[34]. Based on the first detection of BA.1 in South Africa and Botswana, the starting population of infected individuals was defined as follows: Johannesburg, South Africa: 200 infections, 180 mild and 20 severe. Durban, South Africa: 40 infections, 36 mild and four severe. Cape Town, South Africa: 40 infections, 36 mild and four severe. Gaborone, Botswana: 30 infections, 27 mild and three severe.

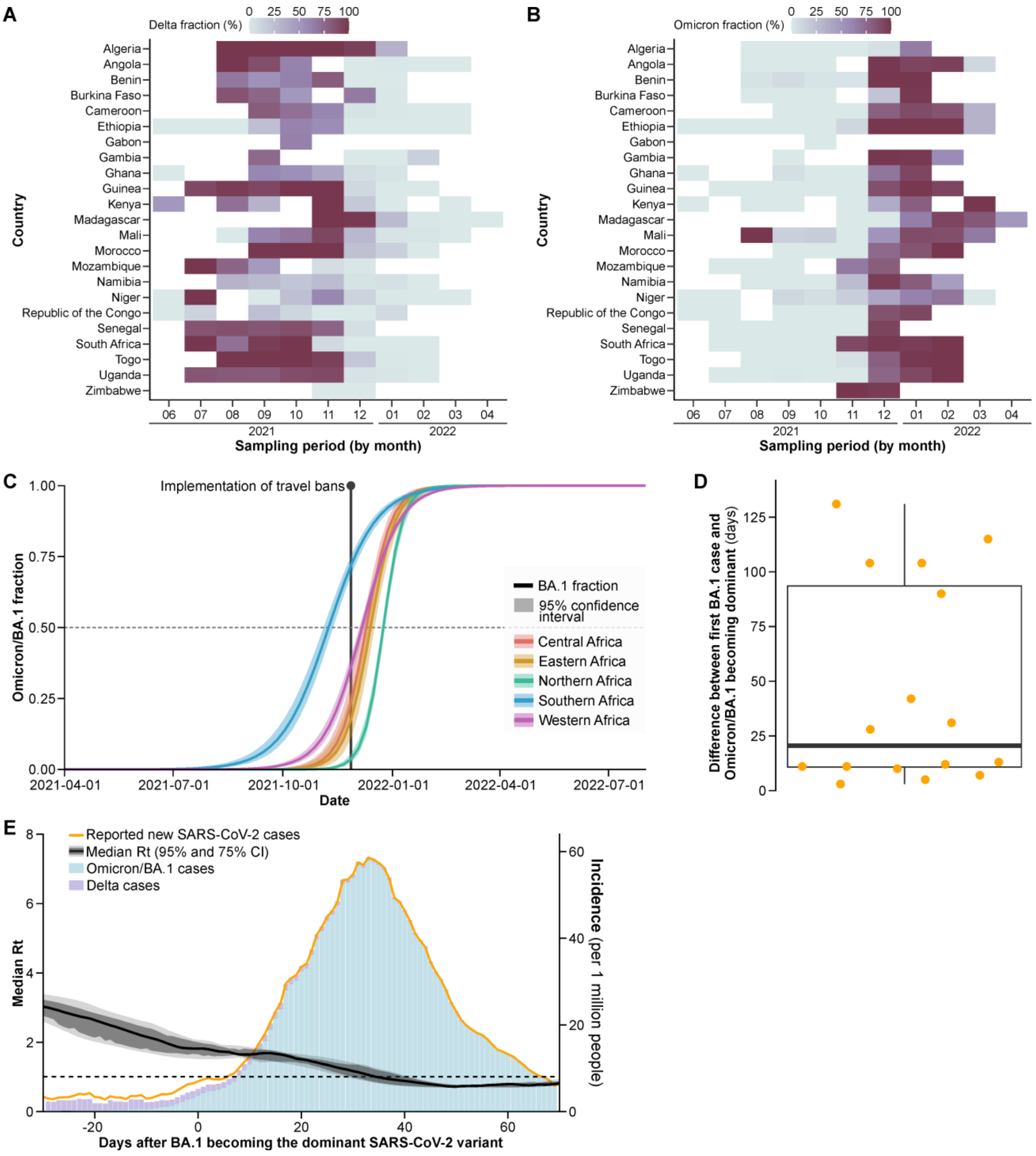


***Figure S4*: Epidemiology of Omicron/BA.1 in Africa not considering travelers.** (**A**) Fraction of samples positive for the Delta marker. (**B**) Fraction positive for the BA.1 marker. (**C**) modelled increase in BA.1 fraction of all SARS-CoV-2 infections per African region based on PCR testing. (**D**) Days until BA.1 became the dominant SARS-CoV-2 variant after its first detection by PCR. (**E**) Smoothed $R_t$ and the incidence among countries represented in this study.

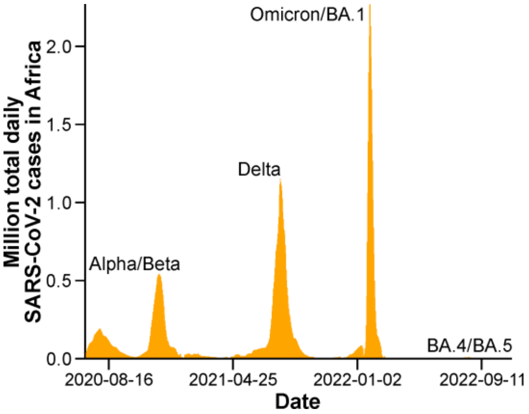


***Figure S5*: Daily reported SARS-CoV-2 cases in Africa by late 2022.**

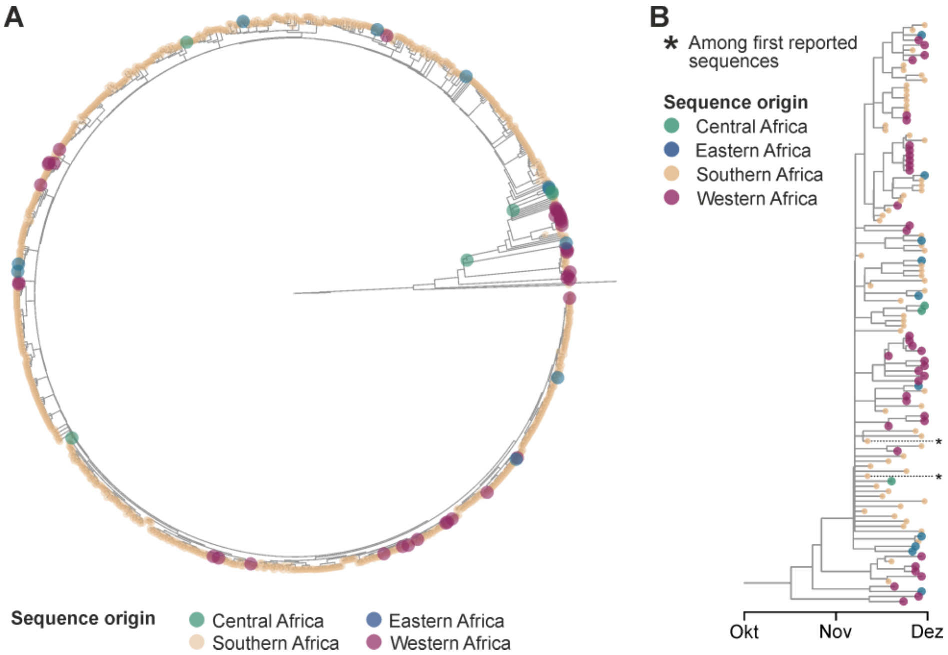


***Figure S6*: Phylogenetic analyses of African BA.1 sequences from human specimens collected before December 2021.** Analyses were conducted using the Nexstrain CLI. Reference sequences are shown by lines only. (**A**) Phylogenetic analyses of all African BA.1 sequences available on GISAID by February 26, 2023. (**B**) Phylogenetic analyses selected sequences. Sequences from Southern Africa were filtered for less than 98% sequence identity using cdhit (https://cd-hit.org). Three sequences from Central Africa with the collection dated before November 21, 2021 were remvoed as their collection date may be false.

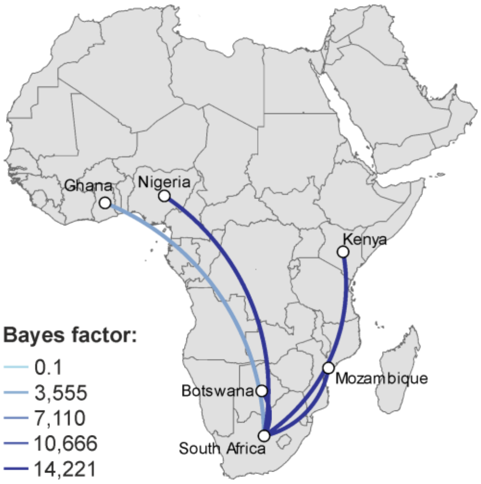


***Figure S7*: Bayes factors for phylogeographic analyses of the BA.1 spread.** Only transitions with a bayes factor > 0.2 are shown for clarity. Three Central African sequences genetically near-identical to BA.1 prototypic sequences (EPI_ISL_17475267, collection date 2021-03-16; EPI_ISL_10023502, collection date 2021-08-12; EPI_ISL_11825829, collection date 2021-10-29) were conservatively removed due to the early collection dates which may be incorrect.

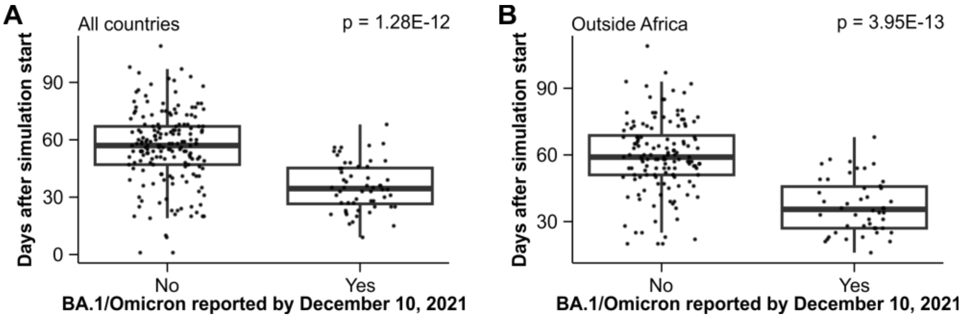


***Figure S8*. Comparison of GLEAMviz simulation and ECDC reports on early BA.1/Omicron cases.** Days after simulation start (November 11, 2021) until 1 BA.1 infection was simulated in 100,000 inhabitants. P-value by Kruskal-Wallis test. (**A**) Comparison among all countries. (**B**) Comparison among non-African countries.

### Supplementary methods

#### Calculation of first expected BA.1 case

The formula used to estimate the time when the first BA.1 case was expected ($T_{1st}$) was:

$$T_{1st} = T_{Dominance} - Log10\left(\frac{0.5}{1/Population}\right)/LOG10(2) * T_{Doubling}$$

$T_{Dominance}$ = Date when BA.1 became dominant (>50% of SARS-CoV-2 cases).

Population = Country-specific population according to United Nations for 2020 (https://population.un.org/wpp/Download/Standard/Population/).

$T_{Doubling}$ = Doubling time.

#### Interpretation of Bayes factors

Bayes factors were interpreted according to Lee MD, Wagenmakers EJ. Bayesian Cognitive Modeling: A Practical Course: Cambridge University Press; 2013.
